# *TUBB4B* variants specifically impact ciliary function, causing a ciliopathic spectrum

**DOI:** 10.1101/2022.10.19.22280748

**Authors:** Sabrina Mechaussier, Daniel O Dodd, Patricia L Yeyati, Fraser McPhie, Thomas Attard, Amelia Shoemark, Deepesh K Gupta, Maimoona A Zariwala, Marie Legendre, Diana Bracht, Julia Wallmeier, Miao Gui, Jacob R Anderson, Mahmoud R Fassad, David A Parry, Peter A Tennant, Alison Meynert, Gabrielle Wheway, Lucas Fares-Taie, Holly A Black, Rana Mitri-Frangieh, Catherine Faucon, Josseline Kaplan, Mitali Patel, Lisa McKie, Roly Megaw, Christos Gatsogiannis, Mai A Mohamed, Stuart Aitken, Philippe Gautier, Finn R Reinholt, Robert A Hirst, Chris O’Callaghan, Ketil Heimdal, Mathieu Bottier, Estelle Escudier, Suzanne Crowley, Maria Descartes, Ethylin W Jabs, Priti Kenia, Jeanne Amiel, Ulrike Blümlein, Andrew Rogers, Jennifer A Wambach, Daniel J Wegner, Anne B Fulton, Margaret Kenna, Margaret Rosenfeld, Ingrid A Holm, Alan Quigley, Diana M Cassidy, Alex von Kriegsheim, Scottish Genomes Partnership, Genomics England Research Consortium, Undiagnosed Diseases Network, Jean-Francois Papon, Laurent Pasquier, Marlène S Murris, James D Chalmers, Clare Hogg, Kenneth Macleod, Don S Urquhart, Stefan Unger, Timothy J Aitman, Serge Amselem, Jean-Michel Rozet, Serge Amselem, Margaret W Leigh, Michael R. Knowles, Heymut Omran, Hannah M Mitchison, Alan Brown, Joseph A Marsh, Julie P I Welburn, Amjad Horani, Jean-Michel Rozet, Isabelle Perrault, Pleasantine Mill

## Abstract

Cilia are small microtubule-based structures found on the surface of most mammalian cells, which have key sensory and sometimes motile functions. Primary ciliary dyskinesia (PCD) is a type of ciliopathy caused by defects in motile cilia. The genetic basis of PCD is only partially understood. Studying a cohort of 11 human patients with PCD, we find that *de novo* mutations in *TUBB4B*, a beta tubulin isotype, cause three distinct classes of ciliopathic disease. *In vivo* studies in mice show that *Tubb4b* plays a specific role in cilia, building centrioles and axonemes in multiciliated cells. Examining the effects of specific TUBB4B variants in cells and in mice, we further demonstrate that distinct *TUBB4B* mutations differentially affect microtubule dynamics and cilia formation in a dominant negative manner. Finally, structure-function studies reveal that different TUBB4B mutations disrupt distinct tubulin interfaces. Importantly, these molecular differences correlate with disease features. We show that tubulin heterodimer-impairing TUBB4B variants underlie nonsyndromic PCD, whilst additional renal and sensorineural ciliopathic features in a syndromic PCD subtype arise from microtubule lumenal interface-impaired TUBB4B variants. These findings suggest that specific tubulin isotypes have distinct and non-redundant subcellular functions, and demonstrate that human tubulinopathies can be drivers of ciliopathic syndromes.

## Introduction

The dynamic remodelling of microtubules drives diverse cellular processes from organelle trafficking and chromosome segregation to templating stable structures such as centrioles and ciliary axonemes. Microtubules are highly conserved polymers composed of alpha and beta tubulin heterodimers. While organisms like *Chlamydomonas reinhardtii* have only single *α*- and *β*-tubulin genes, humans possess 10 *β*-tubulin isotypes, which can pair with one of the 9 isotypes of *α*-tubulin. Single cell sequencing data reveals that tubulin isotypes display both unique and overlapping expression patterns across cell types and developmental stages. Distinct tubulin isotypes, encoded by different genes, provide metazoans with discrete transcriptional modules to meet the demand for tubulin subunits that shift during development and according to cell type (1–3). Coding sequence differences between isotypes within the same species suggest the possibility of functional differences between isotypes that could underlie differences in the physical properties of microtubules into which they are incorporated (4–7). This idea underlies the concept of a ‘tubulin code’ in which expression of a given set of isotypes, combined with specific post-translational modifications, dictates the stability and mechanical properties of the microtubule lattices in which they assemble(3). Whether specific subcellular or organelle-specific lattices exist within a cell remains unclear.

One such subcellular compartment with a distinct microtubule architecture is the cilium. Although different cilia types, in different tissues, may vary in their final structure, function, size and number, they share certain conserved elements. All cilia originate from a basal body, a microtubule-based structure derived from the cell’s centrosome. In the cilium, microtubules are arranged into an axoneme, an axial structure consisting of nine microtubule doublets (MTDs). This arrangement is templated by the basal body, in which microtubules are organized into nine triplets (MTTs). Tubulin subunits are polymerized into protofilaments that are radially interlinked within the MTDs and lengthened longitudinally at the cilia tip. Furthermore, all cilia utilize conserved intraflagellar transport (IFT) machinery to ferry key cargos, including tubulins, along the microtubule axoneme from the cilia base to cilia tip and back. Unlike other cytoskeletal networks in a cell, the microtubules of axonemes are comparatively stable(8, 9) particularly in motile cilia, which require considerable mechanical strength to permit the motor-driven bending necessary to power fluid flow across their surface(10, 11). In motile cilia, these additional ATP-dependent axonemal dynein motors are placed regularly along MTDs as outer and inner dynein arms which drive coordinated cilia beat. Whilst conventional transmission electron microscopy suggests that these microtubule-based structural elements are similar across basal bodies and axonemes, it remains unknown whether axoneme assembly and function utilize specific tubulin isotypes.

Cilia are essential for embryonic development and are also required postnatally, for vision, hearing, smell, respiration, excretion and reproduction. Mutations in over 200 genes that affect cilia structure and/or function result in a growing list of over 40 conditions termed ciliopathies(12, 13). These can be roughly divided into sensory and motile ciliopathies. Sensory ciliopathies result from impaired signaling functions of primary cilia. They are associated with a spectrum of diseases, ranging from lethal multiorgan syndromes to non-syndromic forms like retinal dystrophy, which impact only a specific organ. Motile ciliopathies affect the ability of motile cilia to generate effective fluid flow(14), which can lead to defects in mucociliary clearance and chronic airway disease as well as hydrocephaly due to a build-up of cerebrospinal fluid in the ventricles of the brain. The molecular basis for the clinical heterogeneity observed amongst patients with ciliopathies – even within each group remains unclear.

Primary ciliary dyskinesia (PCD, OMIM: PS244400) is a motile ciliopathy affecting the structure and function of motile cilia lining the airways, the brain ependyma, the reproductive tracts and the transient embryonic node structure involved in left-right patterning during development. In patients with PCD, these cilia are static, beat in an uncoordinated manner, or are completely absent (ciliary agenesis). This can result in chronic respiratory disease, due to impaired mucociliary clearance, as well as laterality defects (e.g., situs inversus), hydrocephaly and infertility in a subset of patients (15, 16). However, syndromic PCD, is very rare(13, 17); PCD patients almost exclusively present with respiratory features with or without involvement of other motile ciliated tissues. The clinical and molecular diagnosis of PCD is challenging. It is a genetically heterogeneous disorder that has significant phenotypic overlap with other more common respiratory diseases. Most cases are recessively inherited, due to variants in 50 genes(18). Mutations in these genes only account for 70% of PCD cases, indicating additional causal genes likely exist.

Here, we report that mutations in the beta tubulin subtype *TUBB4B* are associated with a subgroup of PCD. These mutations impact different functional interfaces of tubulin and result in corresponding distinct presentations of ciliopathic disease, some showing only PCD phenotypes and others exhibiting a syndromic ciliopathy. Through computational, cell-based structure-function analysis, and mouse knockout studies, we define the role of TUBB4B in ciliary function and the effect of disease-associated mutations on microtubule dynamics and axoneme assembly. Furthermore, using engineered disease variants in mice, we demonstrate a distinct dominant-negative disease mechanism. Our study reveals an organelle-specific function for TUBB4B that is not redundant with other tubulin subtypes, and is consistent with the idea of a tubulin code that operates at the subcellular level. These findings also extend our understanding of tubulin-related disorders outside their classical neurodevelopmental and degenerative features, for the first time linking tubulinopathies with the spectrum of ciliopathic phenotypes.

### Identification of *de novo* heterozygous *TUBB4B* variants in PCD cases

To gain further insight into the molecular bases of PCD, we undertook trio whole genome sequencing (WGS) in a cohort of 8 clinically diagnosed PCD patients (Black et al 2022, under revision). We were able to molecularly diagnose 7/8 patients with biallelic variants in known PCD genes. The remaining patient P1 had ciliary agenesis revealed by ultrastructural analysis, sometimes referred to as reduced generation of multiple motile cilia (RGMC), a specific subtype of PCD. This patient also has shunted hydrocephalus. Hydrocephalus in human patients with PCD is rare, but occurs most commonly by recessive inheritance in genes associated with the RGMC phenotype such as *CCNO* and *MCIDAS*, or by heterozygous dominant *de novo* mutations in the master motile ciliogenesis transcriptional regulator *FOXJ1*. However, no pathogenic or potentially pathogenic variants were identified in any of these genes (19–21), or in any other known PCD genes either.

In this patient, however, we identified a *de novo* missense mutation p.P259L (chr9:g.137242994:C>T (hg38)) in *TUBB4B* (**Extended data- Fig 1a**). In collaboration with groups internationally, we identified an additional unrelated ten PCD patients with heterozygous, often recurrent, variants in *TUBB4B*: five patients with p.P259L, one patient with p.P259S (chr9:g.137242993:C>T (hg38)), one patient with an in-frame ten amino acid duplication p.F242_R251dup (chr9: g.137242941_137242970dup (hg38)) and three patients with p.P358S (chr9:g.137243290:C>T (hg38)) (**Fig 1a-c, Extended data- Fig 1**). Common clinical features of airway disease including bronchiectasis were observed across the cohort (**Fig 1d,e**) and in addition 6/11 patients exhibited the less-commonly associated feature of hydrocephaly (**Fig 1f,g, Extended data- Tables 1, 2**). Whilst 8/11 patients presented with PCD only (PCD-only group: p.P259L, p.P259S. p.F242_R251dup), the three patients with the p.P358S substitution all also presented with Leber congenital amaurosis (LCA) associated with sensorineural hearing loss (SNHL), and two out of the three patients also exhibited renal symptoms- a syndromic phenotype suggesting defects in several ciliated tissues (PCD+SND group). These are all distinct from the recurrent TUBB4B missense variants p.R391H or p.R391C previously reported to be responsible for a distinct sensory-neural disease (SND-only) characterized by early-onset and severe retinal dystrophy (EOSRD/LCA) associated with sensorineural hearing loss (SNHL) in 4 unrelated families (22). Importantly, these 4 families report no rhinopulmonary features. This suggests that dominant mutations in TUBB4B cause three clinical presentations: a solely motile ciliopathy (PCD-only), a solely sensory ciliopathy (SND-only) or a novel syndromic form impacting both motile and sensory cilia (PCD+SND).

**Fig. 1.**
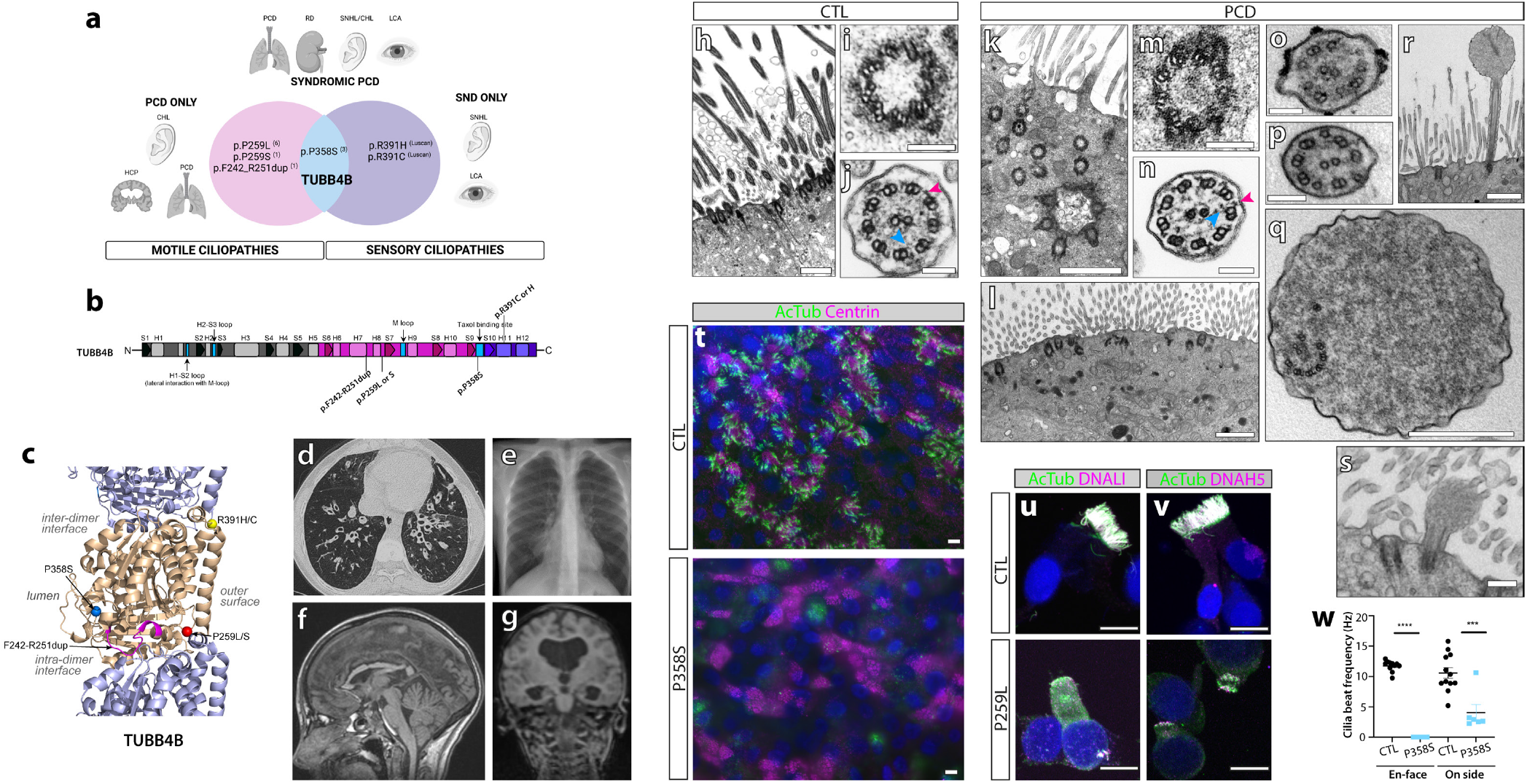
Distinct *de novo TUBB4B* variants cause PCD-only, SND-only or syndromic (PCD+SND) disease. **(a)** Schematic of patient phenotypes clustered on genotypes and where they sit on the ciliopathic spectrum. Number of patients with each variant found in this study or Luscan(22) in superscript in brackets. Abbreviations: CHL: conductive hearing loss; HCP: hydrocephaly; LCA: Leber congenital amaurosis; PCD: primary ciliary dyskinesia; RD: renal disease; SNHL: sensorineural hearing loss; SND: sensory-neural disorder. **(b)** Schematic of protein with resulting changes highlighted below whilst previously reported SND variants are shown above. **(c)** Patient mutations mapped onto an atomic model of TUBB4B (gold) with TUBA1 (purple). **(d-g)** Clinical features of PCD patients: (d) chest CT showing bilateral lower lobe bronchiectasis (P1); (e) X-ray showing right middle lobe atelectasis (P2); (f) midline T1 sagittal image showing irregular corpus callosum secondary to earlier hydrocephaly (shunted), no evidence of basal ganglia dysmorphology is observed, typical of most tubulinopathies (P1); (g) MRI showing dilated ventricles (P8). **(h-s)** Transmission electron microscopy of healthy donor (h-j) and PCD patient nasal epithelia (k-s). Control ciliated epithelia (h), MTTs (i) and cross-section of control axoneme (j). Patient samples show misoriented, internally docked (k) or reduced centrioles without axonemes (l) (P3), incomplete MTTs (P3) (m), and rare intact axoneme (P9) with both inner (blue arrowheads) and outer (magenta arrowheads) dynein arms (n). Other patient axonemal defects include (o) missing doublets (P3), (p) singlet microtubules (P3) or (q) disrupted axonemes (P3). Characteristic (r) rare short axonemes with bulbous tips (r, P3) or (s, P10). **(t)** Wholemount immunofluorescence of nasal epithelial cultures from unaffected mother and patient (P9) (acetylated *α*-tubulin: green cilia; centrin: magenta, centrioles). **(u,v)** Immunofluorescence of healthy donor or patient (P2) cells for cilia axonemes (acetylated *α*-tubulin: green) and dynein motor proteins (q: DNALI: inner dynein arms, magenta or r: DNAH5: outer dynein arms, magenta). **(w)** Cilia beat frequency of control and patient (P9) airway cell cultures. Shown is the mean ± SEM derived from three experimental replicates. ***, p *≤* 0.001; * * **,p *≤*0.0001.

### *TUBB4B* mutations disrupt cilia and centrosomes in patient respiratory cells

Regardless of genotype, strikingly similar cellular phenotypes were observed in PCD *TUBB4B* patient-derived respiratory epithelial cells. These phenotypes include reduced numbers of apically docked basal bodies as well as basal bodies that fail to extend an axoneme (**Fig 1h-l, Extended data- Fig 2a-d**). Axonemes that did extend were short and had bulbous tips displaying disrupted and misoriented microtubules (**Fig 1o-s, Extended data- Fig 2g-j’**). In order to confirm the ciliary agenesis phenotype, control and patient respiratory epithelial cultures were expanded and differentiated. Patient cells in culture displayed poor ciliation and a reduced number of basal bodies (**Fig 1t, Extended data- Fig 2k**), including fewer complete MTTs and apically docked centrioles (**Fig 1i,k,l,r,s**).

**Fig. 2.**
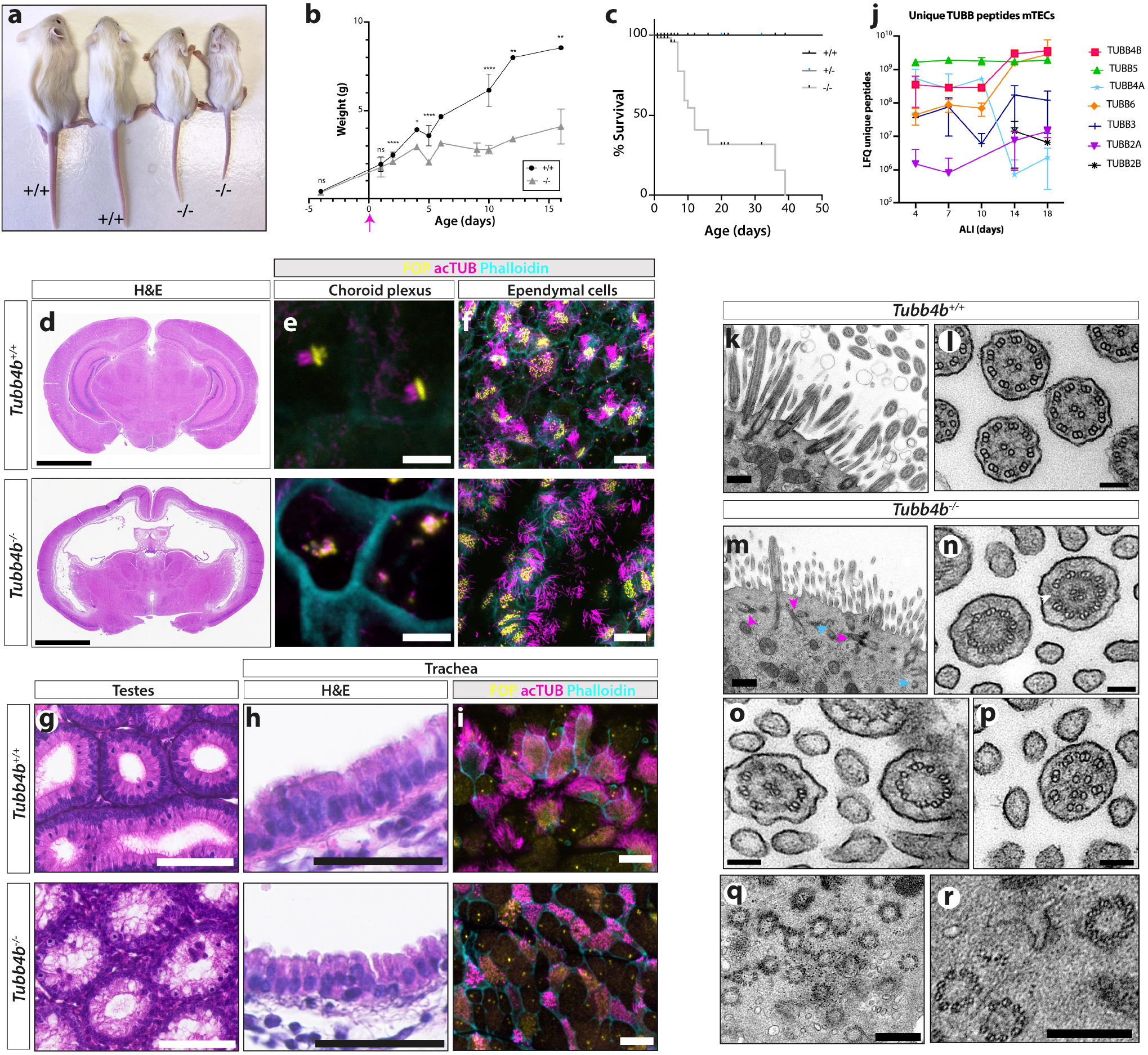
*Tubb4b* is specifically required for axoneme formation in motile ciliated tissues *in vivo*. **(a-c)** *Tubb4b*^*― /―*^ animals exhibit postnatal runting (a) (imaged at P16), starting from P2 (b), and increased postnatal lethality (c) with more than 75% of animals dying before P21. **(d-f)** *Tubb4b*^*― /―*^ animals display hydrocephaly (d) (imaged at P15). Choroid plexus multicilia are profoundly disrupted (e) whilst ependymal cilia appear to have largely normal numbers and lengths (f). **(g-i)** *Tubb4b*^*― /―*^ animals exhibit male infertility with defects in spermatogenesis (g) (imaged at P22) and pronounced absence of cilia in airway epithelia like trachea by histology (h) and whole-mount immunofluorescence (i). **(j)** Mass spectrometry of differentiating wild type mouse tracheal epithelial cell cultures revealed many unique *β*-tubulins are detected. **(k-r)** TEM from control littermate (k,l) and *Tubb4b*^*― /―*^ mutants (m-r) showing misoriented and non-docked centrioles without axonemes (magenta arrowheads), as well as partial centrioles (cyan arrowheads) (m). Mutant *Tubb4b*^*― /―*^ axonemes display missing microtubule doublets (n) (white arrowhead), missing central pair apparatus (o) and microtubule singlets with disrupted organization (p). Within the mutant cytosol, partial centrioles and centrioles with microtubule doublets instead of triplets are common (q, r). (b,c) N= 211 mice. (e, f, i) Fibroblast Growth Factor Receptor 1 Oncogene Partner (FOP): yellow; acetylated *α*-tubulin: magenta; actin: cyan. Scale bars represent: 2.5 mm (d), 5 *μ*m (e), 20 *μ*m (f), 100 *μ*m (g), 50 *μ*m (h), 10 *μ*m (i), 500 nm (k, m, o), 100 nm (l, n) and 300 nm (q, r). (b, j) Graphic bars represent the mean ± SEM derived from N>3 animals per time point (l) and (j) N> 3 biological replicates per time point. Student’s t-test ns, not significant; *, p *≤* 0.1; **, *p ≤* 0.01; * * *, *p ≤* 0.001; * * **, *p ≤* 0.0001.

PCD is most commonly caused by mutations that disrupt the expression and assembly of the axonemal dynein motors powering the cilia beat (23). Immunofluorescence of components of these motors showed mislocalization of the motors either to the cytoplasm or to the apical region of where cilia should form in *TUBB4B* patient cells (**Fig 1u,v**, **Extended data- Fig 2l**). Furthermore, acetylated tubulin, which normally marks axonemes, appears as cytoplasmic aggregates (**Extended data- Fig 2l,m**). These results are consistent with the observed axonemal defects, and suggest that axonemal motors are still produced, even in the absence of axonemes. Indeed, the rare axonemes that did form had inner and outer dynein arms (**Fig 1n**) although with reduced motility (**Fig 1w**). Post-translational modifications of tubulin are a key part of the tubulin code and are common on ciliary microtubules, but patient cells show reduced levels of these marks on the rare cilia observed (**Extended data- Fig 2m**). These defects in centriole amplification, axoneme extension and tubulin modification may underlie the defects in mucociliary clearance observed in patients.

### *TUBB4B* is essential for motile cilia assembly in airway epithelial cells

To investigate the requirement for *TUBB4B in vivo*, we generated *Tubb4b*^*― /―*^ homozygous protein null mice (**Extended data- Fig 3a-c**). *Tubb4b*^*― /―*^ are born at Mendelian ratios (**Fig 2a-c**) but exhibit perinatal lethality with runting (**Fig 2b,c**) and hydrocephaly (**Fig 2d**), both features often associated with motile cilia dysfunction in murine models. We observed defects in surviving male spermatogenesis (**Fig 2g**) and similar defects in the multiciliated cells of the oviduct with reduced cilia numbers and lengths (**Extended data- Fig 3j**). The lack of overt skeletal or growth phenotypes in *Tubb4b*^*― /―*^ neonates suggests that TUBB4B is not required for embryonic development, where primary cilia play key roles. Indeed, *Tubb4b*^*― /―*^ primary cilia in primary fibroblasts show normal numbers and lengths (**Extended data- Fig 3d-f**).

To examine the effects of mutations on motile cilia function, we first evaluated the hydrocephalus phenotype. This phenotype can be caused by defects in motile cilia on ependymal cells, which generate cerebral spinal fluid flow. Given that 75% of the PCD-only cohort of patients also had hydrocephaly, we expected to see defects in the multiciliated ependymal cells lining the ventricles. Mutant animals exhibited pronounced and progressive dilatations of the ventricles neonatally without obstruction of aqueducts, suggesting communicating hydrocephalus (**Fig 2d**). We observed profound reductions in cilia number and length in choroid plexus cells involved in cerebral fluid secretion and regulation (**Fig 2e**). However, motile cilia on ependymal cells had grossly normal lengths and densities (**Fig 2f**). The function of ependymal cilia was further examined *ex vivo* (**Extended data- Fig 3g-i**) and we found no significant difference in ciliary beat patterns or frequency, again with grossly normal cilia numbers and lengths. These data emphasize that, despite the similarities in molecular cascades driving multiciliogenesis between tissue types in mammals, lack of TUBB4B does not cause overt ependymal ciliary defects as it does on the adjacent choroid plexus epithelial cells.

In other tissues, we observed defects in the number and length of *Tubb4b*^*― /―*^ tracheal cilia (**Fig 2h,i**). *Tubb4b*^*― /―*^ centrioles also fail to amplify, and exhibit partially formed MTTs and some docked basal bodies with rare stumpy axonemes (**Fig 2k-r**). These data suggest a unique role for TUBB4B as a critical ‘limiting component’ for organelle size control and scaling in airway epithelial cell cilia.

Strikingly, in the absence of TUBB4B other cytoskeletal processes looked grossly normal including apical-basal patterning in the pseudostratified epithelium. Examination of unique peptides for different tubulin isotypes in total proteomic datasets from control tracheal cultures across differentiation (air-liquid interface day 4 (ALI4): centriole amplification, ALI10: early ciliogenesis) showed that many different *β* tubulin isotypes are expressed during airway epithelial differentiation (**Fig 2j**). However, there is an induction of TUBB4B during early ciliogenesis, whilst its paralogue TUBB4A, different by only 7 amino acids, shows a reciprocal expression pattern. This suggests that while other *β*-tubulins in multiciliated airway cells are expressed across windows where centrioles and cilia are being built, TUBB4B fulfils a unique role in ciliogenesis and is essential for formation of multiple motile cilia in the respiratory epithelium. Thus, TUBB4B is a cilia-specific tubulin.

### TUBB4B variants differentially impact MT dynamics and heterodimer formation

In order to understand how different *TUBB4B* mutations might affect microtubule dynamics and ciliation, we used transient overexpression of human FLAG-tagged proteins in RPE-1 cells. We compared the effects of wild-type and disease-associated variants of TUBB4B (**Fig 3**). PCD-only TUBB4B variants (p.P259L/S) failed to strongly colocalize to microtubules (**Fig 3a,b**) and the PCD+SND syndromic variant (p.P358S) had reduced colocalization. In contrast, microtubule localization was minimally affected for the SND-only variants (p.R391H/C). Under serum-starvation conditions to induce ciliogenesis, we also examined effects of TUBB4B variants on cilia length and numbers (**Fig 3c-e**). We measured the kinetics of microtubule depolymerization in cells expressing these different TUBB4B variants by tracking the number and lengths of microtubules bound to the end-binding protein EB1 after cold shock followed by repolymerization (**Extended data- Fig 4**). PCD-only TUBB4B variants (p.P259L/S), which showed low incorporation into MTs including those of the centrosome, had no observable effects on cytoplasmic MT dynamics (**Fig 3f-h**), but profoundly decreased cilia number and length (**Fig 3c-e**). In contrast, the syndromic PCD+SND TUBB4B variant (p.P358S) localized to centrosomes upon repolymerization but strongly impeded number and length of repolymerizing cytoplasmic MTs, as well as decreasing the number and length of cilia (**Fig 3c-h**). The SND-only variants (p.R391H/C) showed intermediate effects on MT dynamics, and only modestly affected the length of primary cilia (**Fig 3c-h**). Importantly, overexpression of wild-type TUBB4B slightly increased cilia length without disrupting rates of ciliation or microtubule dynamics (**Fig 3c-h**). These findings suggest each of the disease-causing variants acts in a dominant manner but via differing mechanisms to disrupt microtubule biology.

**Fig. 3.**
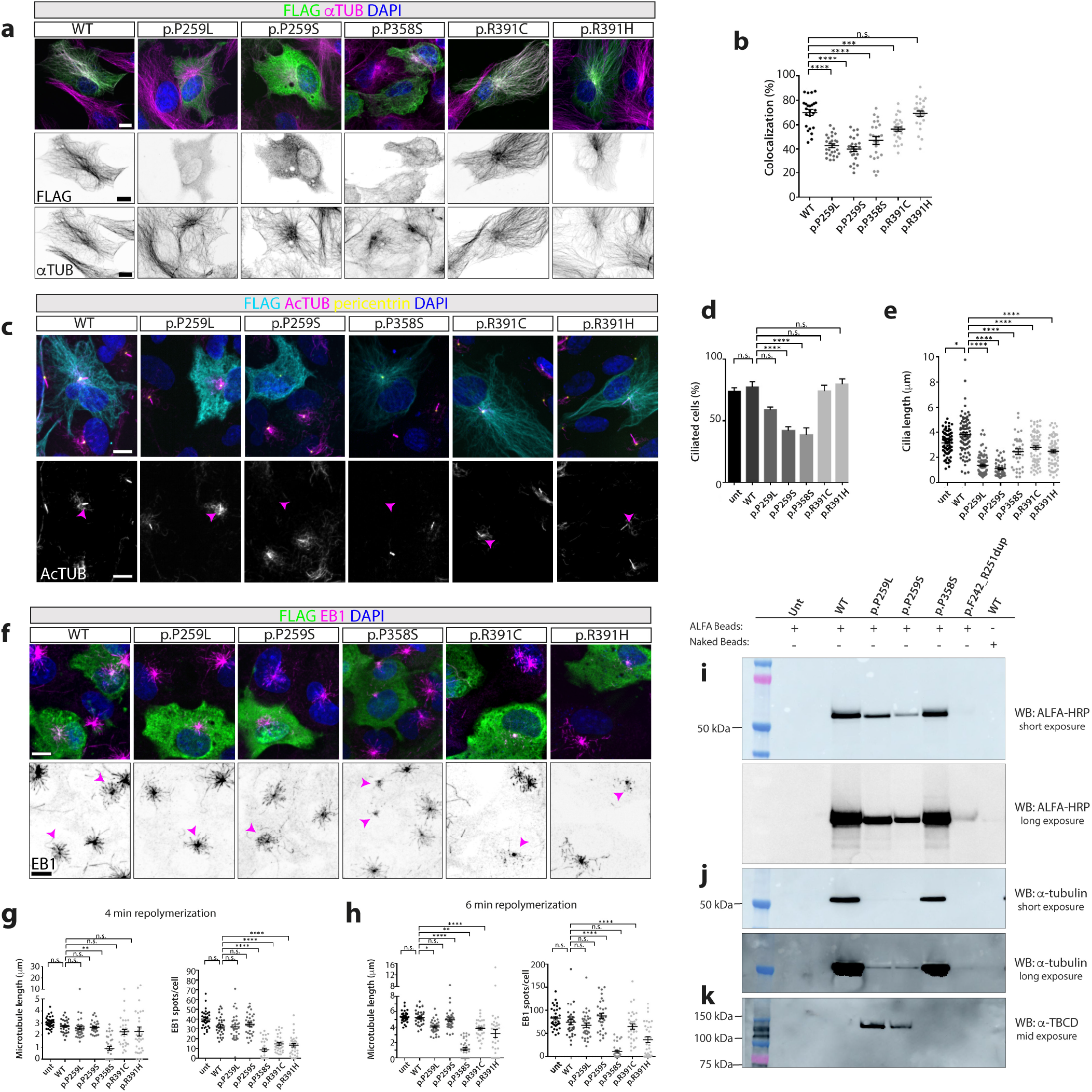
Disease-causing TUBB4B variants alter microtubule dynamics and ciliation. **(a)** Incorporation into the microtubule lattice of RPE1 cells transiently overexpressing wild-type (WT), p.P259L, p.P259S, p.P358S, p.R391C and p.R391H TUBB4B variants, analyzed by immunostaining of FLAG-tagged TUBB4B (green) and *α*-tubulin (magenta) protein in steady-state. DAPI is used to label the nucleus. **(b)** Quantification of percentage colocalization of FLAG and *α*-tubulin staining of RPE1 cells transiently overexpressing TUBB4B variants. **(c-e)** Ciliogenesis of 24 h serum-starved RPE1 cells overexpressing variants, observed by immunostaining of FLAG (cyan), acetylated α-tubulin (magenta) and pericentrin (yellow). DAPI is used to label the nucleus. Acetylated tubulin only channel (lower panel) with transfected cell cilia highlighted by magenta arrowhead. Rates of (d) ciliation and (e) cilia lengths were quantified. **(f)** Microtubule network dynamics analysis of RPE1 cells overexpressing TUBB4B variants, showing immunostaining of FLAG-tagged TUBB4B (green) and EB1 (magenta) protein upon repolymerization at 37 °C for 4 minutes. DAPI is used to label the nucleus. EB1 alone channel (lower panel) with transfected cell highlighted by magenta arrowhead to illustrate variant effects on dynamics. **(g,h)** Quantification upon repolymerization at 4 (g) and 6 (h) minutes. See **Extended data- figure 4** for representative images across cold-induced depolymerization and repolymerization. **(i-k)** Lysates from stable IMCD3 cells expressing ALFA-tagged human TUBB4B patient variants were affinity purified (ALFA beads) and loaded for immunoblot against ALFA (i), *α*-tubulin to examine heterodimer assembly (j) and TBCD, a chaperone involved in tubulin heterodimer assembly pathway (k). Scale bars represent: (a, c, f) 10 *μ*m. Graphic bars represent the mean ± SEM derived from three experimental replicates. ns, not significant; *, p *≤* 0.1; **, p *≤* 0.01; ***, p *≤* 0.001; ****, p *≤* 0.0001.

To gain insight into the molecular mechanisms by which different PCD-causing TUBB4B variants disrupt tubulin heterodimer assembly, we next generated stable cell lines expressing control and PCD-causing TUBB4B variants. While all constructs were equally translated *in vitro* (**Extended data- Fig 5a**), the three PCD-only variants (p.P259L, p.P259S, Dup) in mammalian cells drastically impaired *α*/*β* heterodimer formation. Importantly, p.P358S does not affect binding to *α*-tubulin (**Fig 3i,j**). Moreover, p.P259L and p.P259S also co-purify with TBCD (tubulin folding co-factor D), one of the five co-chaperones required for assembly and disassembly of the *α*/*β* tubulin heterodimers (**Fig 3k, Extended data- Fig 5b**). This suggests these PCD-only variants are stalling during the tubulin biogenesis cycle. We purified in vitro recombinant human wild type TUBB4B and PCD variants co-expressed with human TUBA1A and could confirm that wild type TUBB4B/TUBA1A formed heterodimers robustly in this system (**Extended data- Fig 5c**). The syndromic PCD+SND p.P358S variant appeared relatively stable in this system, with only a moderate reduction in heterodimer formation (**Extended data- Fig 5h,i**), suggesting a mechanism of action downstream of heterodimer assembly. Again the three PCD-only variants disrupted formation of TUBB4B/TUBA1A heterodimers as none could be detected by affinity purification of *α*-tubulin when expressed in insect cells (**Extended data- Fig 5d-g**).

Together this demonstrates how PCD-causing *TUBB4B* mutations disturb centriole number and axoneme size by disrupting heterodimer assembly (PCD-only variants) or disrupting polymerization (PCD+SND) in an organelle-specific manner.

### Mouse and patient cell models demonstrate a dominant negative effect of PCD-associated mutations

To further test the idea that the TUBB4B mutations have a dominant negative effect, we turned to mouse models. If, as proposed, the mutant TUBB4B variants act in a dominant negative manner in vivo, we could expect to see different phenotypes in heterozygotes carrying single patient *Tubb4b* mutations versus null mutations (haploinsufficiency). We therefore used CRISPR-Cas9 mediated genome editing to engineer in mice the *Tubb4b* patient variants carried in PCD-only (p.P259L, p.P259S), syndromic PCD+SND (p.P358S) and SND-only (p.R391H) patients, as well as deletion alleles (**Fig 4a, Extended data- Fig 3a, Extended data- Fig 6a**). We were able to establish the two independent *Tubb4b* null lines from deletion founder animals (**Extended data- Fig 3a-c**), which as heterozygous animals displayed no reduction in airway cilia length (**Extended data- Fig 3d,e**) or fertility defects (**Extended data- Fig 3m**). These two *Tubb4b*^+*/―*^ mouse lines also exhibited normal neonatal survival and growth (**Extended data- Fig 3n**).

**Fig. 4.**
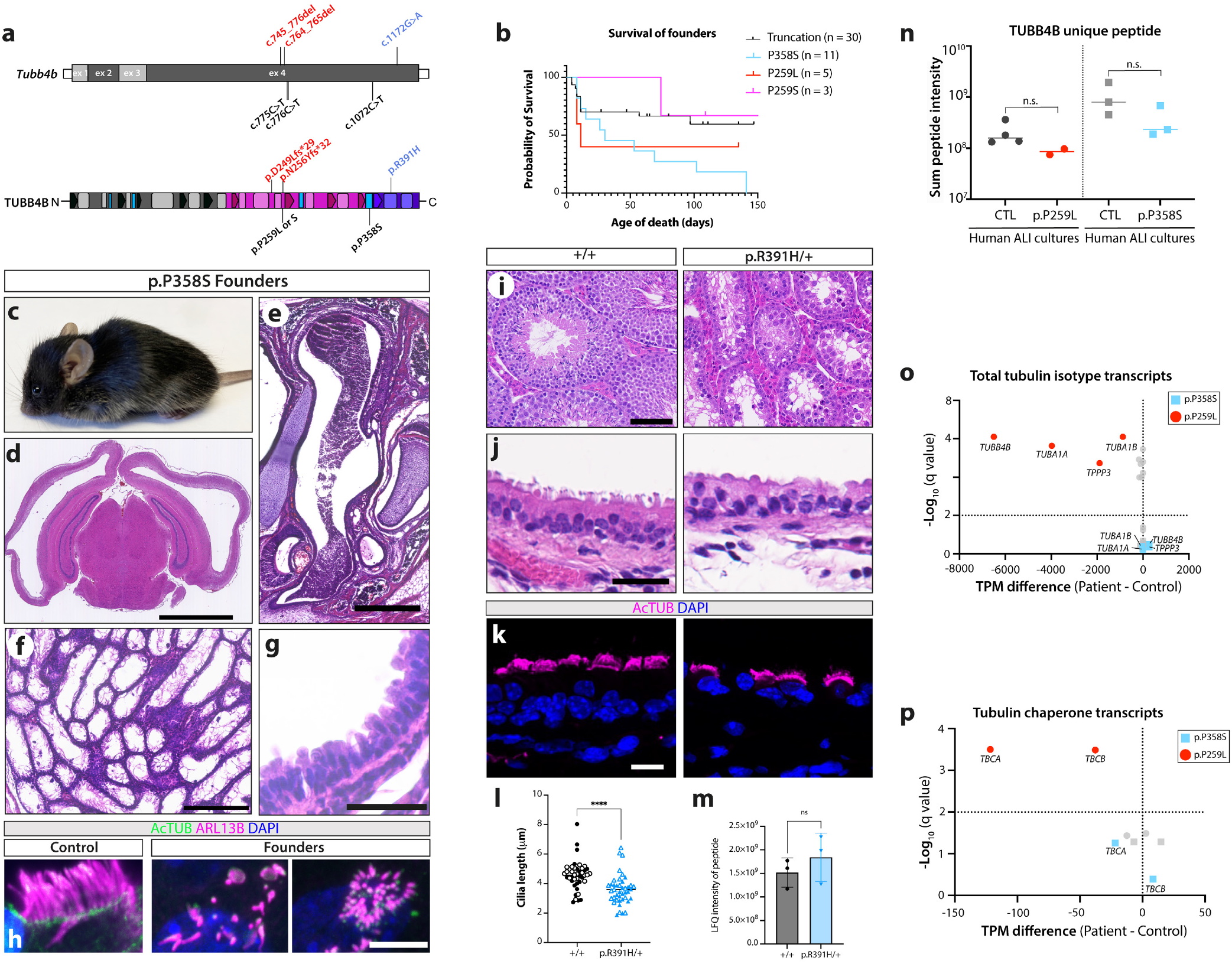
*TUBB4B* variants act via distinct molecular mechanisms and support dominant negative mechanisms of disease *in vivo*. **(a)** Schematic of the human p.P259L/S (PCD-only, red or magenta), p.P358S (syndromic PCD+LCA, blue) and p.R391H (SND-only, purple) mutations and two truncating mutations (p.D249Lfs*29 and p.N256Yfs*32, null alleles (black)) generated by genome editing on the mouse *Tubb4b* mRNA (top) and protein (bottom). **(b-h)** Mice carrying patient variants showed decreased perinatal fitness and survival (b), as well as male and female infertility. (b) Kaplan-Meier graph showing that founder mice carrying patient variants have died either spontaneously or secondary to euthanasia for health concerns. (c-h) p.P358S founder animals exhibited pronounced hydrocephaly (c,d) and mucopurulent nasal plugs (e). Mutant mice showed profound defects in spermatogenesis (f) and loss of tracheal cilia by histology (g) or whole-mount immunofluorescence (h) (acetylated *α*-tubulin: green, ARL13B: magenta). **(i-m)** R391H/+ animals showed no decrease in fitness or survival and females were fertile allowing a line to be established (see also Extended data- figure 5). Males were infertile and showed defects in spermatogenesis (i). Mild defects in the length of cilia on tracheal cells by histology (j) or immunofluorescence (k) (acetylated *α*-tubulin: magenta) (l). Mass spectrometry of trachea extracts quantifying TUBB4B protein levels (m). **(n)** Quantifiecation by mass spectrometry for unique TUBB4B peptides from nasal brush epithelial cultures from healthy controls, a PCD-only patient (red, p. P259L) or syndromic PCD+SND patient (blue, p.P358S). **(o)** RNASeq from nasal brush epithelial cultures of healthy controls or a patient with PCD-only (red, p. P259L) or syndromic PCD+SND (blue, p.P358S) revealed significant upregulation of *TUBB4B, TUBA1A* and *TPPP3* transcripts only in p.P259L patients. **f**. Healthy control human BECs treated the same way as (**e**). (N=9 represents experimental replicates). **(p)** Targeted analysis of RNASeq revealed a significant upregulation in the tubulin chaperone transcripts in PCD-only (red, p. P259L) samples. bars represent: 2.5 mm (d), 500 *μ*m (e), 250 *μ*m (f), 100 *μ*m (i), 50 *μ*m (g), 25 *μ*m (j) and 10 *μ*m (h,k). (l, m) Graphic bars represent the mean ± SEM derived from N=2 biological replicates, n>20 cells/replicate (l) and N= 3 biological replicates. Student’s t-test ns, not significant; ****, p *≤* 0.0001.

Founder mice carrying PCD-causing mutations exhibited increased postnatal lethality compared to founders carrying a range of deletion alleles (**Fig 4b**). They developed pronounced hydrocephaly neonatally **Fig 4c,d**) in addition to defects in mucociliary clearance within the upper airways (**Fig 4e**), with loss of multicilia throughout the respiratory epithelium (**Fig 4g,h**). Moreover, we were unable to transmit any of the PCD variants because surviving founders exhibited both male and female infertility **(Fig 4f**). These mice therefore phenocopy PCD patients.

In contrast, we were able to generate a mouse line carrying the SND-only *Tubb4b* p.R391H variant (**Extended data- Fig 6a,b**), although males remained infertile due to defects in spermatogenesis (**Fig 4i**). *Tubb4b* p.R391H/+ mice did not develop any retinal degeneration (**Extended data- Fig 6c-e**) even when aged, potentially due to differential expression of *Tubb4b* between mouse and human photoreceptors (**Extended data- Fig 6f**). We observed a significant (20%) reduction in airway cilia length (**Fig 4j-l**) in *Tubb4b* p.R391H/+ mice, indicating a dominant effect as we could confirm TUBB4B protein levels were identical between control and p.R391H/+ littermates *in vivo* (**Fig 4m**). However, perhaps because of the different physiology of photoreceptors in mice and humans, SND TUBB4B mutations in mice do not recapitulate the phenotypes of human SND patients. To test this directly in human airway nasal epithelia cultures, we carried out proteomic and transcriptomic analyses on lysates from healthy donors and patients carrying the p.P259L and p.P358S respectively. Due to high conservation between tubulin isotypes, only a single peptide can be used to uniquely identify human TUBB4B in total proteomes. Comparable levels of TUBB4B were detected between controls and both patients (**Fig 4n**), ruling out haploinsufficiency as a disease mechanism. Bulk RNASeq for these two patients and controls revealed distinct molecular signatures. Only the p.P259L patient samples displayed a two-fold increase in *TUBB4B* mRNA (**Fig 4o**) and concomitant increase in mRNAs encoding *TUBA1A* and *TPPP3*, a microtubule polymerizing protein (**Fig 4o**). This is consistent with a tight regulatory control to ensure the appropriate balance of *α* and *β* subunits(24, 25). In keeping with this concept, there is upregulation of the mRNA encoding *TBCA* and *TBCB* the tubulin chaperones involved in binding and stabilizing the nascent *α*-tubulin and *β*-tubulin protein, respectively (**Fig 4p**). These tubulin autoregulation signatures are not observed in the syndromic PCD+SND p.P358S samples, consistent with tubulin heterodimer assembly proceeding normally with this variant (**Fig 3j**).

Together these data indicate that the mode of action of these disease-causing variants is not haploinsufficiency. Despite a decreased intrinsic propensity for the PCD-only variants to assemble into microtubules, transcriptional upregulation of *TUBB4B* itself and its chaperones ensure that some level of mutant protein exists, consistent with a dominant-negative effect. Overall, this suggests a complex series of dominant-negative phenotypes through distinct modes of action on microtubules for each ciliopathy subtype. In the case of mice carrying PCD-causing *Tubb4b* mutations, these models phenocopy many patient features, at both cellular and physiological levels, consistent with acting in a dominant negative manner to disrupt centriolar and ciliary microtubules.

### Different *TUBB4B* variants differentially localize according to clinical phenotype

Although TUBB4B is widely expressed, our mouse knockout studies indicated an essential and non-redundant function for this *β*-tubulin isotype in building airway cilia. In order to understand why, we undertook a structural approach by examining our 3.6 Å resolution cryo-electron microscopy (cryo-EM) structure of the human MTD in axonemes of airway multiciliated cells (26)(**Fig 5a**). We could assign both *α*- and *β*-tubulin isotypes, based on their sidechain density. After evaluating each residue of the candidate isotypes, we determined TUBB4B to be the best fit to the density map and the predominant *β*-tubulin isotype making up airway cilia axonemes in vivo (**Fig 5b**). Thus structural analysis further confirms our findings that TUBB4B is a cilia-specific tubulin, in spite of many other *β*-isotypes being highly expressed within this cell type.

**Fig. 5.**
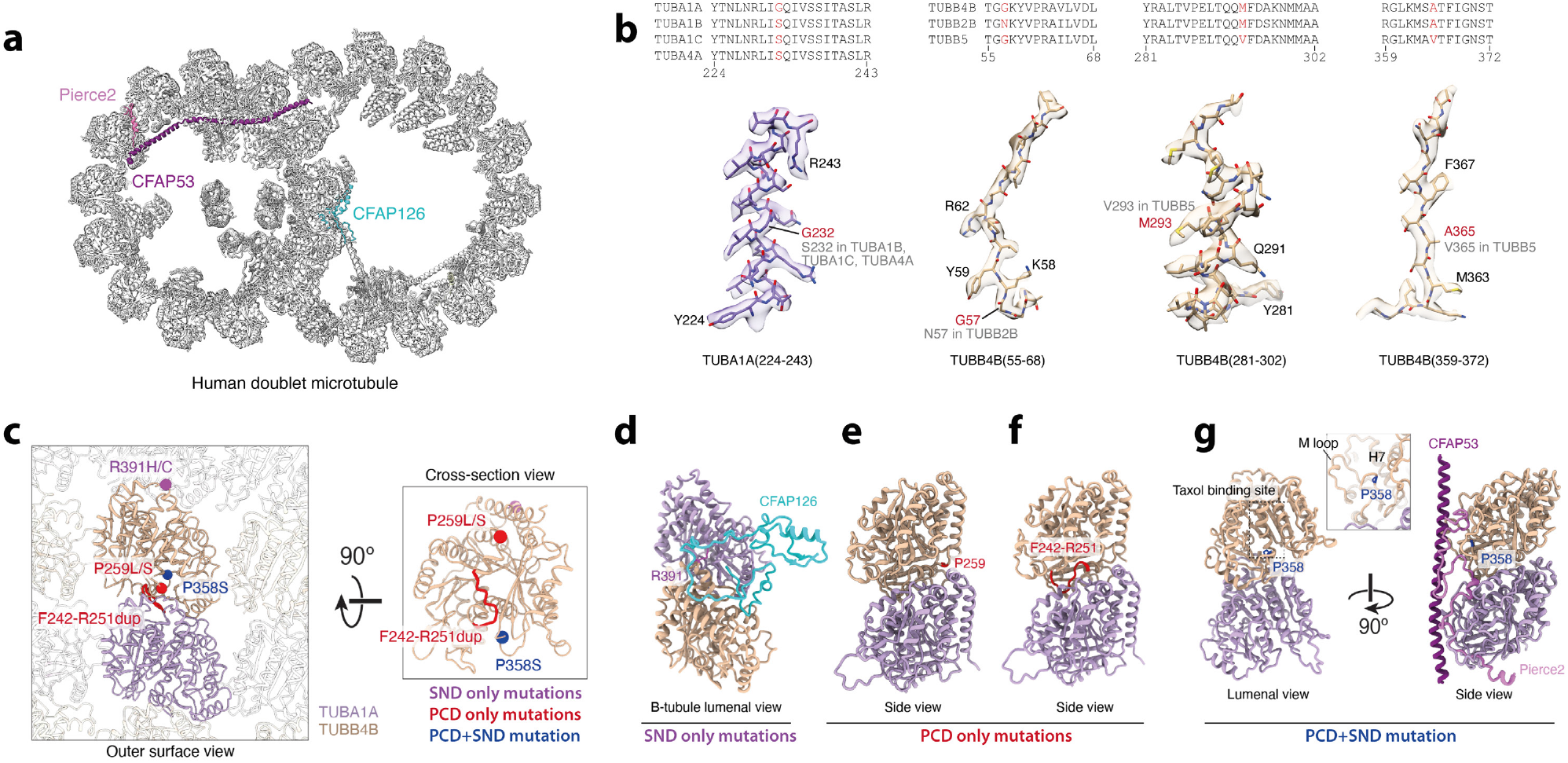
Structural environment of disease-causing variants of TUBB4B. **(a)** Cross section of the human MTD (PDB ID: 7UNG) highlighting proteins that interact with TUBB4B residues associated with disease. The MTD is the conserved cytoskeletal element of both primary and motile cilia, consisting of a complete A-tubule with 13 protofilaments (PFs) and an incomplete B-tubule with 10 PFs. **(b)** The tubulin isotypes that form respiratory axonemal MTDs were determined using sidechain density from the 3.6-Å resolution structure of a human MTD (26). Examples showing how the density corresponding to each site of variation was used to ‘fit’ or to discriminate between candidate residues. After performing this sequence comparison at every variable residue, we determined that the amino acid sequence of TUBB4B was the best fit to the density. **(c)** Orthogonal views showing the positions of disease-causing variants of TUBB4B within the context of the ciliary microtubule doublet (MTD) lattice. The human tubulin isotypes (TUBA1A (purple) and TUBB4B (gold)) were determined based on the cryo-EM density map of human respiratory MTD (26) and their abundance in human multiciliated respiratory cells by single-cell RNA-sequencing(27). Variants are coloured based on their disease association and their positions within the TUBB4B model marked with spheres. Only one TUBB4B molecule is shown in the cross-section view (right). R391 is not visible at this angle. **(d)** Interaction of R391 of TUBB4B with the microtubule inner protein (MIP), CFAP126. **(e-f)** p.P259 and loop p.F242-R251 locate at the intradimer interface. **(g)** p.P358 locates at the taxol binding site which interacts with multiple MIPs including, for example, PIERCE2, as shown on the right.

TUBB4B variants associated to the three different phenotypic classes of disease (SND-only, PCD-only or PCD+SND) were distributed distinctly on the structure of the protein, both within and between tubulin heterodimers (**Fig 5c, Extended data- Table 3**). The previously reported SND-only TUBB4B variants p.R391H and p.R391C (22) localized to the interface between adjacent tubulin heterodimers (**Fig 5d**). The SND p.R391H/C mutations are moderately destabilizing but predicted to more strongly affect longitudinal interactions with the adjacent *α*-tubulin in neighbouring heterodimers. Several other pathogenic missense mutations have been reported in mostly neurodegenerative disorders at this position in other isotypes including TUBB4A p.R391H/L, TUBB3 p.R391L, TUBB2A p.R391H and TUBB8 p.R391C, where these mutations are predicted to disrupt microtubule stability(28, 29).

In contrast, the PCD-only *TUBB4B* variants involve residues not previously associated with human disease. In the PCD-only group (p.P259L/S, p.F242_R251dup) variants localized to the intradimer interface, the interface between each *α*- and *β*-subunit of a tubulin heterodimer (**Fig 5e,f**). Both missense mutations at P259 are predicted to be moderately destabilizing but are more likely to affect the interface with *α*-tubulin (**Extended data- Table 3**). A similar disruption of this intradimer binding interface by the p.F242_R251dup is expected, although the effects of an insertion mutation on protein stability are more challenging to predict.

The PCD+SND syndromic variant (p.P358S) was within the intralumenal face, close to the antitumour drug taxol binding site (**Fig 5g**), where treatment promotes lateral aggregation of taxol-bound protofilaments into stabilized MTs(30). It is also a position known to interact with many microtubule inner proteins (MIPs)(31). The p.P358S mutation is predicted to be moderately destabilizing and it could also disrupt TUBB4B interactions at the intralumenal side of protofilaments, potentially with MIPs or lateral interactions between protofilaments. p.P358L/A/S mutations have also been reported in TUBB8, associated with female infertility(32).

Our combined analysis shows that different *TUBB4B* mutations disrupt distinct molecular aspects of *β*-tubulin and these translate into different ciliopathic disease phenotypes. How these mutations impact tubulin heterodimers and their assembly into higher order structures within cilia and centrioles dictates whether patients present with purely motile ciliopathy features, purely sensory ciliopathy features or a novel syndromic form affecting both cilia types.

## Discussion

Given that cilia are by definition microtubule-based organelles, it is perhaps surprising that mutations in tubulin genes have not previously been reported amongst the ciliopathies. This could suggest a high level of redundancy exists amongst isotypes capable of building cilia. However, our human disease and mouse genetic data indicate that TUBB4B is expressly required for the construction and function of ciliary axonemes. We find that disease-causing TUBB4B variants act in a dominant negative manner to cause a spectrum of ciliopathic diseases. Their locations within the *β*-tubulin molecule create different effects on heterodimer assembly and polymerization into higher order structures of MTDs and MTTs, to impact organelle number and axoneme size. These findings explain the different disease presentations in patients carrying variants affecting distinct *β*-tubulin surfaces.

Differences in patterns and levels of tubulin isotype expression, including TUBB4B, likely explain why certain specialized cilia and tissues are more affected by *TUBB4B* mutations (**Fig 6**). In some axoneme types, like those of MTDs of respiratory cilia (26, 33), we demonstrate that only one predominant *β*-tubulin isotype is utilized. Thus, in these cilia, mutations that inhibit heterodimer assembly completely disrupt centriole biogenesis and axoneme elongation, thus explaining why PCD-only phenotypes are observed; other isotypes cannot compensate. In other tissues, where a ‘mix’ of tubulin isotypes may be used to build different ciliary axonemes, *TUBB4B* variants in PCD-only patients (or in KO mice) do not efficiently integrate into these structures, but other isotypes could compensate and cilia function would not be compromised. Hence, heterodimer-impaired *TUBB4B* mutations result in PCD-only phenotypes. In contrast, the syndromic PCD+SND variant (p.P358S) can robustly integrate into axonemes and thereby disrupt the lattice to cause additional sensory and renal disease phenotypes, in any tissues where TUBB4B is highly expressed. In the polymerization-impaired *TUBB4B* variants found in SND-only patients (p.R391H/C), we observed less dramatic effects on cilia length *in vivo* and *in vitro*. This is consistent with more subtle structural defects on the kinetics or stability of axonemal microtubules into which such variants integrate, and a tissue-specific sensitivity to dysfunction leading to sensory ciliopathy features.

**Fig. 6.**
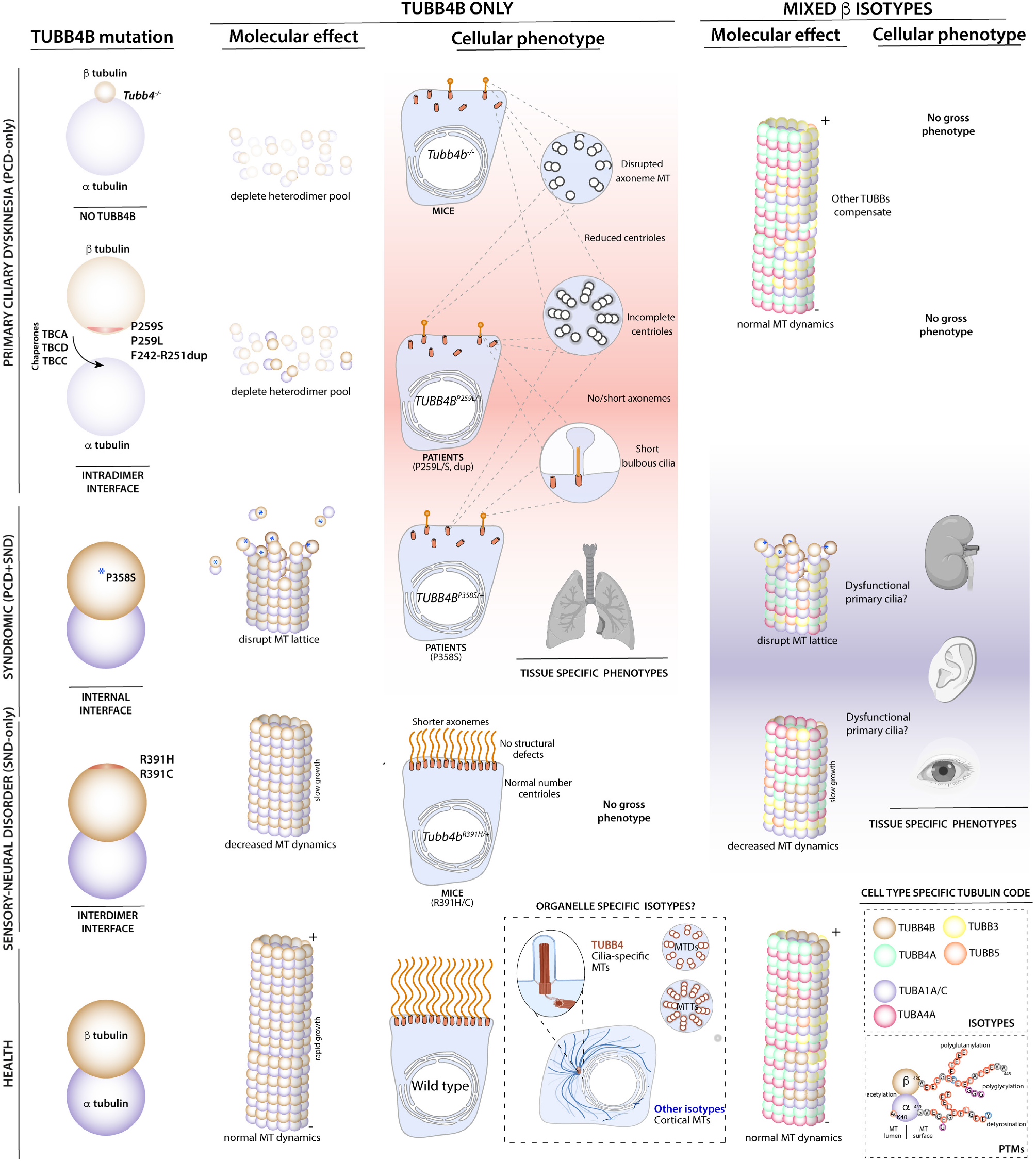
Patient *TUBB4B* variants cause distinct clinical phenotypes depending on how they disrupt tubulin stability and polymerization. A model showing how specific TUBB4B variants confer distinct *de novo*, dominant negative mechanisms of disease. Pathogenic variants which partially destabilize TUBB4B and deplete heterodimer pools phenocopy the null mutationsno other isotype can compensate. They cause PCD-only because in other tissues with other isotypes, other TUBBs compensate. Importantly, although these PCD-only patient variants affect stability, this is not haploinsufficiency as shown by RNASeq and proteomics, as well as a lack of phenotype in heterozygous KO mice. In contrast, P358S which poisons the lattices such that new heterodimers also don’t integrate well, means effective concentration-dependent polymerization during multiciliogenesis also fails causing PCD. However, some P358S TUBB4B also integrates and thus poisons these mixed isotype lattices in other tissues, causing syndromic PCD+SND disease. For the SND-only patient variants, these affect rates of polymerization such that changes in airway cilia length are seen, but animals are not grossly affected. These do affect stability of photoreceptors, likely through the connecting cilium in humans, but not in mice. In short, our patient defined *de novo* mutations have differential effects on tubulin heterodimer stability and polymerization rates, hence affecting different tissues differently, and support a role for TUBB4B as a subcellular specific isotype and a ‘limiting factor’ for building centrioles and cilia on scale.

Within the tubulin code, differences based on expression of isotypes likely underlies some phenotypic differences observed between species. For example, the lack of *Tubb4b* expression in mouse compared with human rod photoreceptors may explain the lack of an eye phenotype in *Tubb4b* p.R391H/+ aged mice (34, 35) (**Extended data- Fig 6c- e**). Expression data suggests that alternate *β*-tubulin isotypes support this role in mice (**Extended data- Fig 6f**). Similarly, whereas fertility in TUBB4B LCA patients appears normal(22), the loss of *TUBB8*, a *TUBB4B* paralogue uniquely expressed during female gametogenesis, leads to reproductive phenotypes (36). Yet mouse *Tubb4b* is expressed in multiciliated cells of the reproductive tract of both males and females, and here fertility defects occur in both male and female *Tubb4b* KO mice (37).

It remains unclear whether different tubulin isotypes are able to compensate for bespoke properties needed to withstand the high mechanical demands of cilia motility and the high structural order of MTDs. In some contexts, such compensation appears likely. In mouse ependymal cells, for example, which express high levels of *Tubb4b* and show a tubulin isotype expression pattern similar to ciliated airway cells (**Extended data- Fig 3o**), centriole amplification and ciliogenesis are nevertheless grossly normal in the absence of TUBB4B. This observation suggests that other isotypes may compensate for the loss of TUBB4B function, including the highly similar TUBB4A(38). These observations are consistent with *Drosophila* studies suggesting that only isotypes with a particular amino acid sequence in the carboxyl terminus (EGEFXXX) are required for normal axonemal function(1, 6, 39, 40). This motif is only found in TUBB4A and 4B in mammals (and TUBB8 in humans), and is the site of heavy post-translational modifications that are associated with cilia stability. These findings raise the interesting possibility that mammalian TUBB4A/B is a motile cilia-specific *β*-tubulin required for the unique mechanical and structural properties of motile axonemes (41). Some of these outstanding questions might be solved by improved spatial proteomics, using strategies such as non-disruptive endogenous tagging of tubulin genes, to determine which tubulin isotypes localize to which microtubule networks across time and cellular space *in vivo*.

Based on our findings, we propose that TUBB4B acts as a regulator of cilia and centriole number and size in multiciliated cells. In this model, local concentrations of *β*- tubulin would be enough to control rates of polymerization into higher order structures, thus acting as a ‘limiting factor’(42, 43). Although tubulin overexpression in isolated axonemes is sufficient to induce ectopic hookor C tubulelike structures (44), we find genetic mutations that effectively deplete pools of heterodimers, through multiple dominant mechanisms, obstruct the rapid polymerization necessary for centriole amplification of M T Ts. We observed both decreased numbers of centrioles as well as incomplete structures. TUBB4B also appears to act as a ‘limiting factor’ for extension of MTDs, given that many docked centrioles fail to form cilia, and those axonemes that do form are shortened and often display bulbous tips as if the cilium has run out of necessary building blocks to template further growth.

In conclusion, our study provides detailed mechanistic insights into how *TUBB4B* variants cause a spectrum of ciliopathic disease that spans both sensory and motile ciliopathies. The disease presentation manifesting in patients depends on how each variant affects tubulin heterodimer pools, as well as the differential composition of the cilia and centrioles into which they incorporate. Our study extends our understanding of tubulinopathies outside of classical neurological features, links them with ciliopathies, and suggests how tubulin diversity in humans underlies and facilitates the diversity of cilia seen *in vivo*.

## Methods

### Subjects

Eleven affected individuals from eleven unrelated families and their healthy relatives were included in the study (5 females and 6 males). Genomic DNA was extracted from peripheral blood by standard procedures. Signed and informed consent was obtained from the affected individual as well as relatives through approved protocols. For P1 (UOE), the study was approved by the London-West London and Gene Therapy Advisory Committee Research Ethics Committee (REC number 11/LO/0883), P2 by London-Bloomsbury Research Ethics Committee (REC 08/H0713/82; IRAS ID 103488), P3 by the Institutional Ethics Review Board of the University Muenster (2015104-f-S), and P4 and P5 by the Institutional Review Board from Institut national de la santé et de la recherche médicale (IRB00003888 -approval n°15-259. Protocols for UNC (P6, P7) human studies were approved by the Institutional Review Board at the University of North Carolina and were performed in compliance with ethical regulations. P8 was recruited for WGS as part of the 100,000 Genomes Project, under approved Research Registry Project RR185 ‘Study of cilia and ciliopathy genes across the 100,000 GP cohort’. For P9, the study protocols were approved by the Institutional Review Board at Washington University in St. Louis. P10 was recruited under studies approved by the Institutional Review Board for Human Use at the University of Alabama at Birmingham (US), in compliance with ethical regulations. P11 was recruited under the Undiagnosed Disease Network protocol 15-HG-0130 approved by the National Institutes of Health Institutional Review Board.

### Whole genome sequencing (WGS) and candidate prioritization

For P1 (UOE), DNA was sequenced by WGS at Edinburgh Genomics (Black et al. 2022 under revision). Libraries were prepared using the Illumina TruSeq PCR-free protocol and sequenced on the Illumina HiSeq X platform. The average yield per sample was 136Gb, with mean coverage of 36x (range 33.9-38.3). After first running analysis with a virtual gene panel of 146 genes, based on the PCD PanelApp panel (v1.14)45 with five additional genes identified in the literature (*CFAP300, DNAH6, DNAJB13, STK36* and *TTC25*), no diagnostic variants were identified in P1. Expanded analysis identified a *de novo* missense mutation p.P259L (chr9:g.137242994:C>T (hg38)) in the gene *TUBB4B* only in the patient, and not present in either parent (**Extended dataFig 1a**). *TUBB4B* is an outlier in gnomAD in terms of constraint, especially in the overrepresented synonymous category (Z-score top 0.2% of all genes, 2x observed vs expected variants). The gene is also highly intolerant to missense variants, (Z-score bottom 0.9% of all genes, 0.24x observed vs expected variants) and intolerant to loss of function variants (probability loss of function intolerant (pLI) 0.81, 0.14x observed vs expected variants). Gene annotation for *TUBB4B* was obtained from gnomAD (v2.1.1, (45) https://gnomad.broadinstitute.org/gene/ENSG00000188229?dataset=gnomad_r2_1) and ExAC (v0.3, (46)). The constraints table was downloaded from the gnomAD website (https://gnomad.broadinstitute.org/). Variant annotation was obtained from the Ensembl Variant Effect Predictor (v99, (47)). We examined the sequence context of P259 residue given it arose repeatedly by independent mutation in 7/11 patients within this study’s cohort. GTCCCGTTT -V–P–F-The change of P>L is CCG>CTG, so deamination of a methylated CpG seems the most likely cause of that mutation. The other change at this residue P>S is CCG>TCG, which could be caused by non-canonical methylation MCG followed by deamination(48). In contrast, the sequence context of P358 which also arose as an independent *de novo* mutation in 3/11 patients is: CCACCTCGG -P–P–R- The change of P>S is CCT>TCT, which is unlikely to be caused by non-canonical methylation. For P8, data was accessed, analysed and filtered as described in (49). Data was reviewed by the Airlock Committee prior to export.

### Whole exome sequencing (WES) and NGS targeted panel

Details of how WES genomic libraries for P2-P7 and P8-P11 were generated, captured and sequenced are summarized in **Extended data – Table 2**.

### Ciliary nasal brushing and high speed videomicroscop

Biopsies and brushing of ciliated epithelium were obtained using a cytology brush from nasal mucosa (inferior turbinate) of the affected individuals P1-P6, P8-P10 and p.R391C and processed for ciliary investigations. All clinical experiments were performed in the absence of acute respiratory tract infections. Cilia beat frequency and pattern were assessed by high-speed video microscopy at a frame rate of >500 frames per second. Video microscopy of ciliated epithelial cells was performed using an inverted microscope with a 20X phase contrast objective (Eclipse Ti-U; Nikon, Melville, NY) enclosed in a customized environmental chamber maintained at 37 °C. Images were captured by a high-speed video camera and processed with the Sisson-Ammons Video Analysis system (Ammons Engineering, Mt. Morris, MI, USA) and analyzed using established methodologies(50). Cilia beat frequency was analyzed in at least 4 fields obtained from each cell preparation. Cells were collected under approval from appropriate local authorities including Washington University institutional Review Board (IRB) approval, the London-Bloomsbury Research Ethics Committee and the local ethics committee DC-2008-512, Paris-Necker.

### Transmission electron microscopy

Airway biopsies were immersed in 2.5% glutaraldehyde and processed by standard procedures for transmission electron microscopy ultrastructural analysis(51). Ultrathin sections were examined at a final magnification of 60000x without knowledge of the clinical data.

### Air liquid interface culturehuman

Primary human nasal epithelial cells were expanded at 37 °C in media selective for basal cells (PneumaCult™-Ex Plus Medium Stemcell™ Technologies, Cambridge, UK) or specialized media51. At 80% confluent basal cells were dissociated and seeded into 6 mm transwell inserts (Corning® Transwell® polyester membrane cell culture inserts, Flintshire, UK). Once a confluent base monolayer had formed, the apical fluid was removed and the basolateral fluid was replaced with ALI media (PneumaCult™-ALI Medium, Stemcell™ Technologies, Cambridge, UK) to promote differentiation. Experiments were performed once cells had been at ALI for at least 3 weeks and were fully differentiated into ciliated epithelium. For RNA and proteomic studies, transwell insert membranes with cells were cut out and stored in RNAlater (ThermoFisher) or snap frozen respectively at -80 °C until use. Cell preparations were maintained in culture for four to 12 weeks. Cells were collected under approval from appropriate local authorities including Washington University institutional Review Board (IRB) approval, the London-Bloomsbury Research Ethics Committee and the local ethics committee DC-2008-512, Paris-Necker.

### Human proteomics studiesin gel digestion

Frozen cell pellets of cultured primary nasal epithelia from healthy unrelated controls or unaffected parent with parallel cultures from patient samples (p.P259L (P1), p.P358S (P9)) were lysed in 2% SDS in PBS and resolved by SDS-PAGE. Each insert was treated as an experimental replicate and graphed separately. Aiming to enrich for tubulin peptides, gel sections (between molecular weight markers of 37-75 kDa) were cut out further dissected into 1 × 1 mm2 fragments. These were dehydrated with acetonitrile (ACN), reduced with 10 mM DTT and 50 mM ammonium bicarbonate (AB) for 20 min at 56 °C, followed by alkylation with 55 mM iodacetamide and 50 mM AB for 1 h RT. Samples were washed by sequential dehydration/hydration steps alternating between ACN and 50 mM AB. Then, samples were digested with trypsin (Promega) at 37 °C for overnight, extracted with 0.1% trifluoroacetic acid (TFA) and 80% ACN 0.1% TFA. The combined eluates were concentrated in a CentriVap Concentrator (Labconco) and loaded onto StageTips. The tryptic peptides eluted from StageTips (80% ACN, 0.1% TFA) were lyophilised and resuspended in 0.1% TFA. Samples were analysed on a Q Exactive plus mass spectrometer connected to an Ultimate Ultra3000 chromatography system (Thermo Scientific, Germany) incorporating an autosampler. 5*μ*L of each tryptic peptide sample was loaded on an Aurora column (IonOptiks, Australia, 250mm length), and separated by an increasing ACN gradient, using a 40min reverse-phase gradient (from 3%–40% ACN) at a flow rate of 400nL/min. The mass spectrometer was operated in positive ion mode with a capillary temperature of 275°C, with a potential of 1,500V applied to the column. Data were acquired with the mass spectrometer operating in automatic data-dependent switching mode, using the following settings: MS 70k resolution in the Orbitrap, MS/MS 17k resolution obtained by HCD fragmentation (26 normalised collision energy). MaxQuant version 1.6. was used for mass spectra analysis and peptide identification via Andromeda search engine (52) using standard settings apart from: Match between runs was enabled. Trypsin or LysC was chosen as a protease with minimum peptide length 7 and maximum of two missed cleavage sites. Carbamidomethyl of cysteine was set as a fixed modification and methionine oxidation and protein N-terminal acetylation as variable modifications. Total proteomic data are available via ProteomeXchange with identifier PXD036304.

### Human transcriptomic studies

Frozen cell pellets of cultured primary nasal epithelia from healthy unrelated controls or unaffected parent with parallel cultures from patient samples (p.P259L (P1), p.P358S (P9)). Briefly, total RNA from each culture was purified using a miRNAeasy Micro Kit (1038703, Qiagen), cleaned up with MinElute (74204, Qiagen) and treated with Turbo DNAse free (AM1907, Invitrogen). RNA quality and integrity were assessed on the Agilent 2100 Electrophoresis Bioanalyser Instrument (G2939AA, Agilent Technologies Inc) using RNA 6000 Nano or Pico (5067-1511, 5067-1513), then quantified using Qubit 2.0 Fluorometer (Q32866, Thermo Fisher Scientific Inc) with a Qubit RNA High Sensitivity assay kit (Q32855). DNA contamination was quantified using the Qubit dsDNA HS assay kit (Q32854). Poly-A mRNA was isolated with magnetic module (E7490, New England Biolabs) from 20-100 ng of each total RNA sample. Libraries were generated with a NEBNEXT Ultra II Directional RNA Library Prep kit (E7760, New England Biolabs) using random hexamers as primers. Sequencing was performed using the NextSeq 500/550 High-Output v2.5 (150 cycle) Kit (20024907, Illumina) on the NextSeq 550 platform (SY-415-1002, Illumina). Total RNASeq data has been deposited in GEO under accession code GSE214070.

### Nasal epithelial cell immunofluorescence

Nasal cells were fixed directly from the patient or after expansion in ALI cultures for wholemount. If direct, samples suspended in cell culture media were spread onto glass slides, air dried, and stored at –80 °C until use. Expanded cultures were fixed directly on the membrane in 4% PFA/PBS, then immunostained and imaged (**Extended data – Table 4**). Nuclei were stained using 4, 6-diamidino-2-phenylindole 1.5 *μ*g/mL.

### Genetic and three-dimensional structure analysis

The 3-dimensional structure of wild-type TUBB4B and mutant variants was predicted using I-TASSER and based on TUBB4B NM_006088.5 reference. Predicted models were aligned on the cryo-electron microscopy structure of a GDP-protofilament (GDP-K MT, EMD-6353, PDB: 3JAS) using the UCSF Chimera software. We modelled the molecular effects of missense mutations using the stability predictor FoldX v5 with all default parameters and three replicates (**Extended data- Table 3**). We used the PDB structure 5FNV, chain B, as it possesses both the inter- and intra-dimeric interfaces with *α*-tubulin. Δ Δ G subunit values represent the predicted effects of mutations on the TUBB4B molecule alone, ignoring any intermolecular interactions, while the Δ Δ G full values were calculated using the full protein complex structure, and thus include effects from the predicted disruption of *α*-tubulin interfaces. To visualise the structure of the Dup variant, we modelled the structure of the full variant protein using SWISS-MODEL.

### Identification of the tubulin isotypes that form axonemal microtubule doublets (MTDs)

The tubulin isotypes that form respiratory axonemal MTDs were determined using sidechain density from the 3.6-Å resolution structure of a human MTD(26). All potential isotypes were first determined by mass spectrometry of the sample used for cryo-EM analysis. Candidates for *α*-tubulin were TUBA1A, TUBA1B, TUBA1C, and TUBA4A. Candidates for *β*-tubulin were TUBB4B, TUBB2B, and TUBB5. Multiple sequence alignments were generated for the *α*and *β*-tubulin isotypes to highlight positions in the primary sequence where the residues differed. The density corresponding to each site of variation was then examined to discriminate between candidate residues. For example, TUBB2B was excluded as an asparagine sidechain at position 57 that does not match the density (**Fig 5b**). The methionine sidechain at position 293 and the alanine sidechain at position 365 of TUBB4B fitted better to the density than the valine (293) and valine (365) of TUBB5, respectively. After performing this sequence comparison at every variable residue, we determined that the amino acid sequence of TUBB4B was the best fit to the density. The same approach was used to identify the *α*-tubulin isotype as TUBA1A, where the glycine of TUBA1A at position 232 was a better fit to the density than the serine of TUBA1B, TUBA1C and TUBA4A. Our assignment is consistent with the abundance of tubulin isotypes in single-cell RNA-sequencing of human multiciliated respiratory cells(53).

### Site-directed mutagenesis

Patient-derived TUBB4B variants c.776C>T, p.P259L; c.775C>T, p.P259S; c.1072C>T, p.P358S; c.1171C>T, p.R391C; and c.1172G>A, p.R391H were generated by mutagenesis via inverse PCR with Phusion polymerase using vector pcDNA3.1-TUBB4B-C-(K)DYK (Genscript, Piscataway, USA) as a respective template with primers listed in **Extended data- Table 4**. The amplified product was digested with DpnI to avoid religation of original non-mutated DNA. Constructs were amplified in XL1-Blue competent cells (Agilent, US), and the whole ORF of each plasmid was Sanger sequenced to confirm the presence of the patient mutation.

### Culture of RPE1 cells

hTERT-RPE1 cells (CRL-4000) were maintained at 37 °C, 5% CO2 in Opti-MEM Glutamax I medium, supplemented with 10% fetal bovine serum and 1% streptomycin/penicillin (Life Technologies, ThermoFisher Scientific). Cells were seeded at 2 × 105 cells/well on glass coverslips in 12-well plates and maintained for 24 hours (80% confluence). Cells were transfected with wild-type or mutant FLAG-tagged TUBB4B plasmids (1 *μ*g, pcDNA3.1-TUBB4B-C-(K)DYK constructs; Genscript) using the FuGene HD transfection reagent according to the manufacturer’s protocol (Promega).

### MT lattice dynamics in RPE1 cells

Microtubule dynamics were characterized 48 h post-transfection. Cells were directly fixed in ice-cold methanol (5 min at -20°C) for the steadystate MT lattice or after having been maintained on ice for 20 and 30 min for MT depolymerization or for 30 min prior to incubation at 37 °C for MT repolymerization for 4 and 6 min., respectively. Fixed cells were permeabilized in PBS supplemented with 3% bovine serum albumin and 0.1% TritonX-100 (1 h RT) prior to immunostaining. Staining colocalization between positive FLAG and positive *α*-tubulin from a given cell area (ROI) was quantified using machine learning of Ilastik software55, percentages of staining colocalization were generated using JACoP plugin on ImageJ software(54) and plotted using GraphPad software. Microtubule lengths were measured by determining the number of EB1 protein spots and the distance between the centrosome and each EB1 signal, in repolymerization state, using a Spot detector plugin within an ROI using Icy software(55). Individual distances were plotted using GraphPad software. Means of fiber alignment degrees, EB1 spot numbers and MT lengths were calculated from two independent experiments (> 30 cells for each cell line). Statistical analyses were carried out by ANOVA and the PLSD Fisher test.

### Ciliary abundance and length in cultured cells

Transfected cells were propagated for 24 h at 37 °C, 5% CO2 in serumfree Opti-MEM Glutamax I medium to promote ciliation. Cells were fixed in ice-cold methanol and immunostained. Mean numbers of ciliated cells and cilia lengths were calculated from two independent experiments (> 100 cells for each condition) using ImageJ(54). Statistical analyses were carried out by the PLSD Fisher test according to the significance of the Student’s t test.

### IMCD3 TUBB4B stability and interaction studies

Human TUBB4B (NM_001372.3) constructs (wild type, p.P259L, p.P259S, p.P358S and Phe242_Arg251dup) were ordered from GeneWiz, Germany with a C-terminal ALFA tag using the sequence 5’ –AGCAGGCTGGAGGAGGAGCTGAGGAGGAGGCTGAC CGAGTAG-3’. A single proline linker 5’-CCC-3’ was included between the cDNA and tag. Plasmids were sub-cloned into the pCDH-CMV-MSC-EF1*α*Hygro plasmid (CD515B-1; Systems Biosciences, USA), using Nhe1 and Not1 restriction enzymes. Lentiviruses with a VSV-G pseudotype were produced (Viral Vectors Core, Shared University Research Facilities, University of Edinburgh, UK). To generate stable IMCD3 cell lines expressing control or patient variants, IMCD3 cells (CRL-2123, ATCC) were transduced with 3 × 10*∧* 7 copies/mL of virus with polybrene (H9268, Sigma) final concentration of 10 *μ*g/mL in DMEM-F12 (12634010, Gibco), 10% FCS, 1% P/S to 80% confluency. After 24 hours, fresh media was added. After 96 hours following transduction, hygromycin was added to the media at a final concentration of 100 *μ*g/mL and cells were selected for 7 days. To test the ability of TUBB4B variants to heterodimerize, IMCD3 stable cell lines expressing wild type and patient variants fused in frame with the small terminal tag ALFA (NanoTag Biotechnologies) were grown to 80% confluency. Plates were placed on ice for 30 min to depolymerise microtubules after which the culture media was aspirated and cells scraped into 400 *μ*l BRB80 buffer (80 mM PIPES, 1 mM MgCl2, 1 mM EGTA, pH 6.8) plus 10% glycerol, 0.2% Triton X-100, 5 μg /ml DNase I, Halt Protease inhibitor (Pierce) and 1 mM GTP (R1461, Thermo Scientific). Cells were lysed in a water bath sonicator for 10 min, incubated at 37 °C for 20 min and centrifuged at 13K rpm. Cleared supernatants were used to determine total protein levels or incubated with 20 *μ*l ALFA SelectorPE beads (N1510, NanoTag Biotechnologies, Germany) for affinity capture of TUBB4BALFA for 1 hour at RT, washed x 4 times with BRB80 buffer and 10% Glycerol. Bound proteins were released by competition with 0.1 mg of ALFA-elution peptide (N1520, NanoTag Biotechnologies, Germany) for 15 min RT. Resin eluted or total lysates were resolved in acrylamide gels and transferred using Trans-Blot® Turbo™ Transfer System (170-4150, Biorad) with transfer reagents Trans-Blot TurboTM (Biorad 170-4270), then followed by iBindTM (SLF1000, Thermo Scientific) and iBindTM Flex Solutions (SLF2020, Thermo Scientific). Blots were immunoblotted using antibodies listed in Extended data- Table 4 and imaged following incubation with chemiluminescent substrate SuperSignalTM West Pico Plus (34580, Thermo Scientific) on the ImageQuant 800 (Amersham) using either auto-exposure or manual with indicator of saturation. All quantifications done in non-saturated bands using ImageJ.

### In vitro transcription and translation studies

Plasmids (pCDH-CMV-MSC-EF1*α*-Hygro) containing the human TUBB4B (NM_001372.3) wild type and variants (p.P259L, p.P259S, p.P358S and Phe242_Arg251dup) with a Cterminal ALFA tag were used for TNT expression with the TnT ® T7 Quick Coupled Transcription/Translation System (L1170, Promega), according to the manufacturer’s protocol. Reactions were loaded into NuPAGE precast gels, transferred onto PVDF membrane (1704272, BioRad), and then rinsed in water then TBST, and then blocked in 4% BSA in TBS plus 0.1% Tween. Membranes were then incubated overnight 4 °C with ALFA-HRP (N1502-HRP, NanoTag Biotechnologies, Germany) in 5% milk TBST then washed 3 × 10 min TBST and developed using Pierce SuperSignal Pico Plus (Pierce) reagent and imaged on ImageQuant.

### In vitro studies: Cloning, protein expression and purification

TUBB4B (NM_001372.3) wild type and mutants were subcloned into the pFastbac Dual vector containing His6TUBA1 and TUBB3-FLAG, where they replaced TUBB358. A sequence encoding a PreScission protease-cleavable FLAG tag (LEVLFQGPGGSGGDYKDDDDK) was added to the Cterminus of TUBB4B. The Bac-to-Bac System (Life Technologies, Inc.) was used to generate bacmids for baculovirus protein expression. SF9 cells were grown to a density of 2 × 106 cells/mL and infected with viruses at the multiplicity of infection of 1. Cultures were grown in suspension for 48 hours in Sf-900 III SFM (Thermo Fisher), and cell pellets were collected, washed in PBS, and frozen at -80 °C. Cells were lysed through 3 freeze-thaw cycles in 1x BRB80 buffer (80 mM PIPES, pH 6.9, 1 mM MgCl2, 1 mM EGTA) with addition of: 0.5 mM ATP, 0.5 mM GTP, 1 mM PMSF, and 25 units/μl benzonase nuclease. The lysate was supplemented with 500 mM KCl and cleared by centrifugation (30 min at 20,000 x g). To study the TUBB4B mutants, His6-TUBA1 was purified using Ni2+-NTA–agarose beads (GE Healthcare Life Sciences) according to the manufacturer’s guidelines. The supernatant was mixed with a nickel-nitrilotriacetic acid resin (Qiagen) equilibrated with high salt buffer (BRB80, 40 mM imidazole). His-tagged tubulin was eluted with 250 mM imidazole in BRB80 buffer. For purification of FLAG-tagged TUBB4B, the eluate was purified on anti-FLAG G1 affinity resin (GenScript). FLAG-tagged tubulin was eluted by incubation with FLAG peptide (GenScript) at 0.25 g/L concentration. Porcine brain tubulin was purified as described59 and stored in liquid nitrogen. Tubulin heterodimers were further purified using size-exclusion chromatography pre-equilibrated in gel filtration buffer (BRB80 supplemented with 0.1 mM GTP). Western blotting was performed according to standard protocols with details of antibodies in **Extended data- Table 4**.

### Generation of mouse lines and patient mutation F0 founder mouse models

For the *Tubb4b*^*R*391*H/*+^ line, the NIH guide was followed for the care and use of laboratory animals, with approval from the French Ministry of Research (APAFiS 20324) and following ethical principles in the LEAT Facility of Imagine Institute. For all other lines, animals were maintained in SPF environment and studies carried out in accordance with the guidance issued by the Medical Research Council in “Responsibility in the Use of Animals in Medical Research” (July 1993) and licensed by the Home Office under the Animals (Scientific Procedures) Act 1986 under project license number P18921CDE in facilities at the University of Edinburgh (PEL 60/6025). Generation of *Tubb4b*^*R*391*H/*+^ mice using CRISPR/Cas9 as described in **Extended data- Figure 6**, using guides detailed in **Extended data- Table 4**. To generate *Tubb4b*^+*/―*^ mice (*Tubb4b*^*KO*1*/*+^ and *Tubb4b*^*KO*2*/*+^) we used CRISPR/Cas9 as described in **Extended data- Fig 3**, using guides detailed in **Extended data- Table 4**

To generate animals *Tubb4b*^*P* 259*L/*+^, *Tubb4b*^*P* 259*S/*+^ and *Tubb4b*^*P* 358*S/*+^ mice, a similar targeting strategy was attempted using a variety of guide and repairs detailed in **Extended data- Table 4**. Silent mutations were included to block the re-cutting as reported to increase the HDR accuracy and efficiency(56). However, we saw increased perinatal lethality and infertility in founders carrying patient-derived mutations and no lines could be established even using IVF techniques. Briefly, C57BL/6J female mice were super-ovulated and fertilized embryos were injected at the 1 cell stage. The microinjection mix consisting of RNPs with 0.35 *μ*M of guides and 1.8 *μ*M of recombinant GeneArt Platinum Cas9 (Thermo Fisher Scientific, US) and 20 ng/*μ*l ssODN repair templates (Integrated DNA Technologies (IDT), US) was incubated at 37 °C for 10 min prior to pronuclear microinjection. Zygotes were cultured overnight before transfer to pseudopregnant CD1 females. Founders were genotyped by PCR and Sanger sequenced from genomic DNA, isolated from ear biopsies, using primers (see **Extended data- Table 4**). To establish colonies of *Tubb4b*^*R*391*H/*+^ and *Tubb4b*^+*/―*^ mice, founder mice were crossed with C57BL/6J and CD1 mice respectively to remove potential off-targets, and the heterozygous offspring then outcrossed to C57BL/6J and CD1 respectively at least 5 generations to maintain a colony. CD1 was used for *Tubb4b*^+*/―*^ mice to reduce the severity of neonatal lethality and hydrocephaly, coupled to small litter size, characteristic of C57BL/6J mice. Genotyping was performed using primers detailed in **Extended data- Table 4** followed by Sanger sequencing in house or by Transnetyx (Cordova, TN).

### Electroretinographic analysis of mouse models

The function of the retina of ten *Tubb4b*^*R*391*H/*+^ and ten wild-type C57BL/6J mice, aged of 2 months to 1 year, was analysed by electroretinography using the CELERIS Next Generation Rodent ERG Testing platform, according to the manufacturer protocol (Diagnosys LLC, Cambridge, UK) and guidelines for animal safety. In brief, mice were dark-adapted overnight, anesthetized according to their weight and exposed to two four-step sequences of increasing light stimuli from 0.01 to 3 cd.s/m2 and two-step sequences of 3 to 10 cd.s/m2, to elicit and record rod and cone-specific responses, respectively. Statistical analyses were carried out using GraphPad software using the post hoc Sidak’s test (two-way ANOVA).

### Mouse trachea proteomics

Tracheas from mutant and wild type littermate mice aged between P1 and P5 for Tubb4b KO experiments and between P40 and P100 for Tubb4b391His/+ experiments were used for total proteomics, using a filter aided sample preparation method (FASP) method. Tracheas were flash frozen in liquid nitrogen and stored at -70 °C. Samples were lysed in 2% SDS, 0.1 M DTT, 0.1 M Tris-HCl pH 7.6 by pipetting, heating to 95 °C for 3 min and then removing non-solubilized material. FASP purification and double digestion was done following the protocol described in61 with following alterations. We used Vivaspin500 30k cut-off ultrafiltration devices (Sartorius) and 50 *μ*L aliquots. All other steps, including washes, reduction and alkylation were identical. The resulting sample was diluted to 50 *μ*L with 0.1 M Tris-HCl pH 8.5 and 1 *μ*g of endoproteinase LysC (Wako) was added. After overnight incubation at 37 °C, the LysC fraction was collected by centrifugation of the filter units for 20 min. The sample in the filter was then diluted with 50 *μ*L of 0.1 M Tris-HCl pH 8.5 containing 1 *μ*g of trypsin. Following a 4 h digestion, tryptic peptides were collected by centrifugation of the filter units for 20 min. Samples were acidified with 1% trifluoroacetic acid (TFA) and desalted using StageTips, dried using a CentriVap Concentrator (Labconco) and resuspended in 15 *μ*l 0.1% TFA. Protein concentration was determined by absorption at 280 nm on a Nanodrop 1000, then 2 *μ*g of de-salted peptides were loaded onto a 50 cm emitter packed with 1.9 μm ReproSil-Pur 200 C18-AQ (Dr Maisch, Germany) using a RSLC-nano uHPLC systems connected to a Fusion Lumos mass spectrometer (both Thermo, UK). Peptides were separated by a 140-min linear gradient from 5% to 30% acetonitrile, 0.5% acetic acid. The Lumos was operated using following settings: MS 120k resolution in the Orbitrap, MS/MS obtained by HCD fragmentation (30 normalised collision energy), read out in the ion-trap with “rapid” resolution with a cycle time of 1s. The Limma package was used for mass spectra analysis and peptide identification(57). Total proteomic data are available via ProteomeXchange with identifier PXD036304.

For **Figure 2j**, we specifically analysed unique peptides for all alpha and beta tubulins from the total proteomes from control mTECs across differentiation time points we previously generated(27). Briefly, here total mTEC proteomes were derived from two animals/genotype with three experimental replicates per time point (days 4–10, animal pair 1; days 14–18 animal pair two). The data were analyzed using the MaxQuant 1.6 software suite (https://www.maxquant.org/) by searching against the murine Uniprot database with the standard settings enabling LFQ determination and matching. The data were further analyzed using the Perseus software suite. LFQ values were normalized, 0-values were imputed using a normal distribution and standard settings.

### Histology and immunohistochemistry of mouse tissues

Mouse tissues were obtained at different stages after euthanasia by cervical dislocation, anaesthetic overdose or CO2 asphyxiation, performed according to protocol guidelines for animal safety. Upon dissection, tracheas, kidneys and brains were fixed in 4% PFA/PBS, testes fixed in Bouin’s fixative, and eyes were fixed in Davidson’s fixative according to standard protocols. Tissues were serially dehydrated and embedded in paraffin. Samples were sectioned at 5-8 μm and processed for hematoxylin-eosin (HE) using standard protocols. Wild-type C57BL/6J mice were used as a reference in all knock-in *Tubb4b*^*R*391*H/*+^ analyses. Paraffin embedded eye and trachea tissue sections were were dewaxed and rehydrated via ethanol series prior to antigen retrieval in 10 mM Tris-HCl pH 9.2, 2 mM EDTA, 0.01% Tween-20 for 7 min at 900 W in the microwave. Sections were blocked for 1 h with blocking solution (0.1% Tween, 50 mM NH4Cl, 1% BSA and PBS) prior to immunostaining using primary and secondary antibodies detailed in **Extended data- Table 4**. Samples were stained with 1.25 *μ*g/mL DAPI (Roche, Mannheim, Germany), rinsed and mounted in Fluoromount medium (Sigma) under glass coverslips.

### Whole-mount immunofluorescence

Tracheas, choroid plexuses and oviducts from littermates or age matched controls were dissected in PBS and fixed in 4% methanol-free formaldehyde for 1 h RT. Samples were permeabilised in PBS/0.5% Triton-X-100 for 15 min and blocked in 4% BSA/PBS/0.025% Tween-20 (PBST). The corresponding antibody incubations were done overnight in 4% BSA/PBST (**Extended data- Table 4**). Following washes and staining with (DAPI), samples were mounted in Prolong Gold (Life Technologies, Thermo Fisher Scientific).

Brain ventricles were dissected according to 64, pre-extracted with 0.1% Triton X in PBS for 1 minute, then fixed in 4% PFA or ice cold methanol for at least 24 h at 4 °C, followed by permeabilization in PBS (0.5% Triton X-100) for 20 min room temperature. Ventricles were blocked in 4% BSA in PBST (PBS/0.25% Triton X-100) for 1 h at room temperature, then placed ependymal layer down in primary antibodies (**Extended data- Table 4** in 4% BSA/PBST for at least 12 h. Ventricles were washed in PBS 3 × 10 min and incubated with secondary antibodies (**Extended data- Table 4**) in 4% BSA/ PBST (0.25% Triton X-100) at 4 °C for at least 12 h. Ventricles were washed in PBS 3 × 10 min, and mounted on glass bottom dishes (Nest, 801002) in Vectashield (VectorLabs), immobilized with a cell strainer (Greiner Bio-One, 542040).

### Isolation and immunofluorescence of primary mouse cells

Mouse tracheal epithelial cells (mTECs) were isolated and cultured as described previously(58, 59). Ependymal cells were isolated from mice aged P0 to P5 and cultured as described previously(60). Mouse fibroblasts were harvested from a mix of tail and ear tissue from P5 mice as described(61). For immunofluorescence, cells were plated on coverslips or in glass bottom plates. Ependymal and fibroblast cells were fixed in 4% methanol-free formaldehyde for 5-10 min. Samples were permeabilised in TBST (TBS/0.1% Triton-X-100) for 5 min, blocked in 5% donkey serum in TBST. The corresponding primary and secondary antibody incubations (**Extended data- Table 4**) were done overnight in 1% donkey serum in TBST. Washes were done in TBST and stained with DAPI before mounting in Prolong Gold (Life Technologies, Thermo Fisher Scientific).

### Single cell RNASeq analysis

Published 10X single cell data and metadata was read using the Seurat(62–65) SCTransform method to obtain a gene by cell expression matrix using data from(66–68) or the published gene by cell matrix was used with data from(34, 35). The proportion of cells of each of the cell types, as identified in the data by the respective authors, expressing a tubulin (expression greater than zero) was calculated as a proportion of all cells of that type. The resulting matrix was visualised as a heatmap allowing rows (cell types) to cluster.

### Imaging

Images for Fig 1t were acquired using epifluorescent microscopy. Brightfield images were captured on a Hamamatsu Nanozoomer XR (Hamamatsu Photonics, Japan) with 20x and 40x objectives. Confocal Z-stack projections in **Fig 1u,v**, **Fig 3, Extended data- Fig 2k, Fig 4k** and **Extended data- Fig 6d** were captured on a Spinning Disk Zeiss microscope (Zeiss, Oberkochen, Germany). Confocal Z-stack projections in **Extended data- Fig 2l,m, Fig 4h** and **Fig 2e,f,i** were taken on a Nikon A1+ Confocal (Nikon Europe B.V., Netherlands) with Oil 60x or 100x objectives with 405, Argon 561 and588, 640 lasers and GaSP detectors. 3D reconstructions of images in **Extended data- Fig 3f,i,j** were captured with an Andor Dragonfly and Mosaic Spinning Disc confocal using Nikon oil 40X or 100X lenses. High speed video microscopy for mouse samples was performed on a Nikon Ti microscope with a 100X SR HP Apo TIRF Objective, and Prime BSI, A19B204007 camera, imaged at 250 fps. Projections, 3D reconstructions or panels were generated using ImageJ (National Institutes of Health), NIS Elements (Nikon Europe B.V., Netherlands), NDP.view2 (Hamamatsu Photonics, Japan) or Imaris software 9.9 (Oxford Instruments, UK). Final composite images were generated using FigureJ plugin on ImageJ software (National Institutes of Health, Bethesda, MA, USA)(69), Photoshop or Illustrator (Adobe Systems, San Jose, CA).

### Data analysis

Data analysis was carried out in Microsoft Excel, GraphPad Prism 9 (version 9.4.1, GraphPad Software, USA) and Matlab. Statistical tests are described in the figure legends.

## Supporting information

Mechaussier Dodd 2022 medRXiv extended data files

## Data Availability

All data produced in the present work are contained in the manuscript, with details of datasets listed in the Methods section. Total proteomic data are available via ProteomeXchange with identifier PXD036304. Total RNASeq data has been deposited in GEO under accession code GSE214070.

https://www.ncbi.nlm.nih.gov/geo/query/acc.cgi?acc=GSE214070,

## Acknowledgements

We are grateful to all of the patients and families for their participation in the study. We thank Dr Lee Murphy and colleagues at the Wellcome Trust Clinical Research Facility; Edinburgh Genomics for sequencing and analysis; and the Brompton Hospital PCD Diagnostic Service and South East Scotland Genetics Service for clinical support. We are grateful to the IGC Advanced Imaging Resource, the IGC Mass Spectrometry facility and HGU Bioinformatic Analysis Core, as well as the Newcastle University Electron Microscopy Research services, and, in particular, Tracey Davey for outstanding support on this project. We thank investigators and coordinators of the Genetic Disorders of Mucociliary Clearance Consortium that is part of the Rare Disease Clinical Research Network, Dr. Jaclyn Stonebraker and Elizabeth Schecterman for technical assistance, Dr. Hong Dang for Bioinformatics help, and Kelli Sullivan and Nicole Capps for help with specimen accrual and coordination. We thank Dr. Shrikant Mane, Dr. Weilai Dong and Dr. Francesc Lopez-Giraldez from the Yale Center for Mendelian Genomics (UM1 HG006504) for providing whole exome sequencing and bioinformatics support. This research was made possible through access to the data and findings generated by the 100,000 Genomes Project. The 100,000 Genomes Project is managed by Genomics England Limited (a wholly owned company of the Department of Health and Social Care). The 100,000 Genomes Project is funded by the National Institute for Health Research and NHS England. The Wellcome Trust, Cancer Research UK and the Medical Research Council have also funded research infrastructure. The 100,000 Genomes Project uses data provided by patients and collected by the National Health Service as part of their care and support. We thank Pierre David at the Transgenesis platform LEAT at the Imagine Institute, Dr Romain Luscan, Nicolas Goudin at Necker BioImage Analysis, SFR Necker and Dr Aminata Touré at Institut Cochin for help and support with this project.

