## Supplementary material for "*TUBB4B* variants specifically impact ciliary function, causing a ciliopathic spectrum": Mechaussier Dodd 2022 medRXiv extended data files

[1]Laboratory of Genetics in Ophthalmology, INSERM UMR 1163, Institute of Genetic Diseases,Imagine, Paris University, Paris, France, 75015
[2]MRC Human Genetics Unit, Institute of Genetics and Cancer, University of Edinburgh, Edinburgh, UK, EH4 2XU
[3]Wellcome Trust Centre for Cell Biology, School of Biological Sciences, University of Edinburgh, Edinburgh, UK, EH9 3BF
[4]Respiratory Research Group, Molecular and Cellular Medicine, University of Dundee, Dundee, UK, DD1 9SY
[5]Royal Brompton Hospital, London, UK, SW3 6NP
[6]Department of Pediatrics, Washington University School of Medicine, St. Louis, MO, USA,63110
[7]Department of Pathology and Laboratory Medicine, Marsico Lung Institute, University of North Carolina at Chapel Hill, NC, USA,2759
[8]Molecular Genetics Laboratory, Sorbonne Université, Assistance Publique - Hôpitaux de Paris (AP-HP), Hôpital Armand Trousseau, Paris, France, 75012
[9]Sorbonne Université, INSERM, Childhood Genetic Disorders, Paris, France,75012
[10]Department of General Pediatrics, University Children's Hospital Münster, Münster, Germany, 48149
[11]Department of Biological Chemistry and Molecular Pharmacology, Blavatnik Institute, Harvard Medical School, Boston, MA, USA, 02115
[12]Genetics and Genomic Medicine Department, UCL Institute of Child Health, University College London, UK, WC1N 1EH
[13]Department of Human Genetics, Medical Research Institute, Alexandria University, Alexandria, Egypt,
[14]Faculty of Medicine, University of Southampton, Southampton, UK, SO17 1BJ
[15]Centre for Genomic and Experimental Medicine, MRC Institute of Genetics and Cancer, University of Edinburgh, Edinburgh, UK, EH4 2XU
[16]South East of Scotland Genetics Service, Western General Hospital, Edinburgh, UK, EH4 2XU
[17]Department of Anatomy, Cytology and Pathology, Hôpital Intercommuncal de Créteil, Créteil, France, 94000
[18]Biomechanics and Respiratory Apparatus, IMRB, U955 INSERM – Université Paris Est Créteil, CNRS ERL 7000, Créteil, France, 94010
[19]MRC Prion Unit at UCL, UCL Institute of Prion Diseases, London, UK, W1W 7FF
[20]Center for Soft Nanoscience and Institute of Medical Physics and Biophysics, Münster, Germany, D-48149
[21]Biochemistry Division, Chemistry Department, Faculty of Science, Zagazig University, Ash Sharqiyah, Egypt
[22]Core Facility for Electron Microscopy, Department of Pathology, Oslo University Hospital-Rikshospitalet, Oslo, Norway, 20 0372
[23]Centre for PCD Diagnosis and Research, Department of Respiratory Sciences, University of Leicester, UK
[24]Department of Medical Genetics, Oslo University Hospital, Oslo, Norway, 0450
[25]Paediatric Department of Allergy and Lung Diseases, Oslo University Hospital, Oslo, Norway, 0450
[26]Department of Genetics, University of Alabama at Birmingham, Birmingham, AL, USA, 35233
[27]Icahn School of Medicine at Mount Sinai, New York, NY, USA, 10029
[28]Department of Paediatric Respiratory Medicine, Birmingham Women's and Children's Hospital NHS Foundation Trust, Birmingham, UK
[29]Département de Génétique, Hôpital Necker-Enfants Malades, Assistance Publique Hôpitaux de Paris (AP-HP), Paris, France, 75015
[30]Laboratory of Embryology and Genetics of Human Malformations, INSERM UMR 1163, Institut Imagine, Université de Paris, Paris, France, 75015
[31]Carl-Thiem-Klinikum Cottbus, Cottbus, Germany, 03048
[32]Department of Ophthalmology, Boston Children's Hospital, Boston, MA, USA, 02115
[33]Department of Otolaryngology, Boston Children's Hospital, Boston, MA, USA, 02115
[34]Department of Pediatrics, University of Washington School of Medicine and Seattle Children's Research Institute, Seattle, WA, USA
[35]Division of Genetics and Genomics and the Manton Center for Orphan Diseases Research, Boston Children's Hospital, Boston, MA, USA
[36]Department of Paediatric Radiology, Royal Hospital for Children and Young People, Edinburgh, UK, EH16 4TJ
[37]Cancer Research UK Edinburgh Centre, Institute of Genetics and Cancer, University of Edinburgh, Edinburgh, UK, EH4 2XU
[38]Department of Pediatrics, Harvard Medical School, Boston, MA, USA
[39]Genomics England, London, UK. William Harvey Research Institute, Queen Mary University of London, London, UK, EC1M 6BQ
[40]Undiagnosed Diseases Network Coordinating Center, Harvard Medical School, Boston, MA, USA
[41]ENT Department, Bicêtre Hospital, Assistance Publique-Hôpitaux de Paris (AP-HP), Paris-Saclay University, Le Kremlin-Bicêtre, France, 94270
[42]Medical Genetics Department, CHU Pontchaillou, Rennes, France
[43]Department of Pulmonology, Transplantation, and Cystic Fibrosis Centre, Larrey Hospital, Toulouse, France
[44]Department of Paediatric Respiratory and Sleep Medicine, Royal Hospital for Children and Young People, Edinburgh, UK, EH16 4TJ
[45]Department of Child Life and Health, University of Edinburgh, Edinburgh, UK, EH16 4TJ
[46]Department of Pediatrics, Marsico Lung Institute, University of North Carolina at Chapel Hill, NC, USA, 27599-7020
[47]Department of Medicine, Marsico Lung Institute, University of North Carolina at Chapel Hill, NC, USA,27599-7020
[48]Department of Cell Biology and Physiology, Washington University School of Medicine, St. Louis, MO, USA, 63110
[*]These authors contributed equally to this work



**tubulin | cilia | primary ciliary dyskinesia | ciliopathies | tubulinopathies | axonemes | centrioles| cytoskeleton | disease modelling | microtubules**



| | | | | Primary Ciliary Dyskinesia (PCD) | | | | | | | | | | | | Sensorineuronal disease (SND) | | Mutation |
|---|---|---|---|---|---|---|---|---|---|---|---|---|---|---|---|---|---|---|
| Case number | Samples recruited | Sex | Age range at diagnosis | Ciliary ultrastructure | Cilia motility | nNO (ppb) | Current FEV1 (% predicted) | NRD | Chronic wet cough | Regular IV antibiotics | Recurrent infections | Rhinosinusitis | Bronchiectasis | Recurrent ear infection | Hydrocephaly | Auditory Defects | ERG | HGVS protein |
| P1 | Trio | F | 11-15 | Ciliary agenesis | Abnormal | 15.5 | 47 | YES | YES | YES | YES | YES | YES | NO | YES | CHL | Normal | p.P259L |
| P2 | Trio | F | 0-5 | Ciliary agenesis | NA | 50 | 107 | YES | YES | NO | YES | YES | YES | NO | NO | SNHL/CHL | NA | |
| P3 | Trio | F | 0-5 | Ciliary agenesis | NA | NA | NA | YES | YES | NA | YES | YES | NA | YES | YES | Hearing deficit | NA | |
| P4 | Family | M | 6-10 | Ciliary agenesis | Abnormal | 23 | 76 | YES | YES | YES | YES | YES | YES | YES | YES | SNHL/CHL | NA | |
| P5 | Singleton | F | 0-5 | Ciliary agenesis | NA | 35 | NA | NO | NA | NA | YES | YES | YES | NO | YES | NA | NA | |
| P6 | Family | F | 6-10 | Ciliary agenesis | NA | 6.9 | 45 | YES | YES | YES | YES | NA | YES | YES | YES | CHL | NA | |
| P7 | Singleton | M | 11-15 | NA | NA | NA | NA | YES | NO | YES | YES | NA | YES | YES | NO | SNHL | NA | p.F242_R251dup |
| P8 | Duo | M | 0-5 | Ciliary agenesis | Abnormal | 6 | 75 | YES | YES | YES | YES | YES | YES | NO | YES | CHL | Normal | p.P259S |
| P9 | Trio | M | 0-5 | Ciliary agenesis | Abnormal | 6.7 | NA | YES | YES | NO | YES | YES | NO | YES | NO | SNHL/CHL | Flat | p.P358S |
| P10 | Trio | M | 11-15 | Ciliary agenesis | Abnormal | NA | NA | NA | NA | NA | YES | YES | NA | NA | NA | SNHL | Flat | |
| P11 | Family | M | 6-10 | NA | NA | NA | NA | YES | NA | NA | NA | NA | NA | YES | NO | SNHL | Flat | |

**Extended data- Figure 1: Clinical features of *TUBB4B* cohort reported in this study.**

| Case number | Change (hg38) | HGVS cDNA | HGVS protein | Zygosity | Inheritance Mode | ACMG Criteria | Platform used |
|---|---|---|---|---|---|---|---|
| P1 | chr9:137242994:C:T | c.776C>T | p.P259L | Heterozygous | de novo | PS2-PM2-PM6-PP2-PP3 | WGS: Illumina HiSeq X |
| P2 | chr9:137242994:C:T | c.776C>T | p.P259L | Heterozygous | NA | PM2--PP2-PP3 | Capture: Agilent SureSelectQXT NGS kit<br>WES: Illumina NextSeq |
| P3 | chr9:137242994:C:T | c.776C>T | p.P259Leu | Heterozygous | de novo | PS2-PM2-PM6-PP2-PP3 | Capture: Agilent SureSelect All Exon V.6<br>WES: Illumina HiSeq2500 |
| P4 | chr9:137242994:C:T | c.776C>T | p.P259L | Heterozygous | de novo | PS2-PM2-PM6-PP2-PP3 | Capture: Roche MedExome capture<br>WES: Illumina NextSeq |
| P5 | chr9:137242994:C:T | c.776C>T | p.P259L | Heterozygous | NA | PM2-PP2-PP3 | Capture: Roche SeqCap EZ Choice capture<br>WES: Illumina MiSeq |
| P6 | chr9:137242994:C:T | c.776C>T | p.P259L | Heterozygous | de novo | PM2-PM6-PP2-PP3 | Capture: Roche xGen Exome v1.0<br>WES: Illumina Novaseq 6000 |
| P7 | chr9:137242934:G:GCCTGCGCTTCCCAGGCCAGCTCAATGCTGA | c.723_752dup | p.F242_R251dup | Heterozygous | NA | PM2-PM4 | Capture: Roche xGen Exome v1.0<br>WES: Illumina Novaseq 6000 |
| P8 | chr9:137242993:C:T | c.775C>T | p.P259S | Heterozygous | NA | PM2-PP2-PP3 | WGS: Illumina HiSeq 2500 |
| P9 | chr9:137243290:C:T | c.1072C>T | p.P358S | Heterozygous | de novo | PS2-PM2-PP2-PP3 | WES: GeneDx |
| P10 | chr9:137243290:C:T | c.1072C>T | p.P358S | Heterozygous | de novo | PS2-PM2-PP2-PP3 | Capture: Agilent SureSelect All Exon V5<br>WES: Illumina HiSeq2500 HT |
| P11 | chr9:137243290:C:T | c.1072C>T | p.P358S | Heterozygous | de novo | PS2-PM2-PM6-PP2-PP3 | WES: GeneDx WGS: Baylor Genetics |

### Extended data- Figure 2: Genetic features of patient variants in *TUBB4B*.

| Case numbers | Change (hg38) | HGVS cDNA | HGVS protein | PolyPhen2 HumVar score | SIFT score | Grantham score | ΔΔG subunit (kcal/mol) | ΔΔG full (kcal/mol) | Study |
|---|---|---|---|---|---|---|---|---|---|
| P1-P6 | chr9:137242994:C:T | c.776C>T | p.P259L | probably_damaging (0.993) | deleterious_low_confidence(0.02) | moderately conservative (98) | 1.44 | 4.6 | This study |
| P8 | chr9:137242993:C:T | c.775C>T | p.P259S | possibly_damaging (0.847) | deleterious_low_confidence(0.01) | moderately conservative (74) | 3.14 | 4.92 | |
| P9-P11 | chr9:137243290:C:T | c.1072C>T | p.P358S | possibly_damaging (0.551) | deleterious_low_confidence(0.04) | moderately conservative (74) | 2.31 | 2.22 | |
| Family 1-3 | chr9:140137842G>A | c.1172G>A | p.R391H | probably damaging (0.993) | deleterious (0) | conservative (29) | 0.99 | 3.41 | Luscan et al 2017 |
| Family 4 | chr9:140137841C>T | c.1171C>T | p.R391C | probably_damaging (0.999) | deleterious (0) | radical (180) | 1.5 | 2.77 | |

### Extended data- Figure 3: Description of patient variants in *TUBB4B* and predicted pathogenicity.

Standard predictors struggle to predict pathogenicity of tubulin variants accurately. Using structural modelling of variant effects on both the $\beta$-tubulin monomer (subunit) and its interaction with $\alpha$-tubulin a heterodimer (full) based on the crystal structure, we could demonstrate profound effects on heterodimer formation in PCD-only variants. Here, higher $\Delta\Delta$G means the variant is predicted to destabilize the protein and importantly its interactions (full). Interactions, like lateral interactions between subunits not in the structure, are not captured for interfaces affected like p.P358S.

### Extended data- Figure 4: Segregation and location of pathogenic variants in *TUBB4B* identified in unrelated PCD patients.

**(a)** P1 was recruited as a trio for WGS based genetic diagnosis of PCD (Black et al. 2022 under revision). Expanded analysis however identified a heterozygous *de novo* missense mutation p.P259L (chr9:g.137242994:C>T (hg38)) in *TUBB4B* (NM_006088.6) that was present only in the patient. Mutation and the de novo pattern of segregation of the allele were confirmed by targeted Sanger sequencing. **(b)** P2 is proband of a family initially screened for 34 PCD genes including *CCNO* and *MCIDAS* by high-throughput sequencing, followed by screening of a UCL Great Ormond Street ICH targeted gene panel of 40 PCD and 400 motile cilia genes. Whilst biallelic mutations in *DNAH9* were identified comprising a splice acceptor mutation (NM_001372.3:c.7553-3del) and exonic missense mutation (NM_001372.3:c.12640G>T, p.Gly4214Cys), the cellular phenotype of reduced generation of motile cilia was not in keeping with pathogenic variants in this gene(70). Reanalysis of sequencing data confirmed a heterozygous *TUBB4B* de novo missense mutation p.P259L (chr9:137242994:C>T (hg38)) only in the patient, and not present in either parent. **(c)** P3 is proband from a trio recruited for WES based genetic diagnosis of PCD, followed by targeted analysis of variants in *TUBB4B* (NM_006088.6) which identified a heterozygous *'de novo'* missense variant [c.776C>T; p.P259L] in exon 4 in the patient that was not present in either parent or a non-related control. **(d)** P4 is a proband recruited as a family for WES genetic diagnosis of PCD, which identified a heterozygous de novo missense mutation p.P259L (g.chr9:137242994:C>T (hg38)) in *TUBB4B*. Mutation confirmation and segregation of the allele were performed by targeted analysis of the *TUBB4B* locus using Sanger sequencing. **(e)** P5 is proband recruited as a singleton for genetic diagnosis of PCD. After a negative targeted capture sequencing of 50 PCD genes, Sanger sequencing of *TUBB4B* identified a heterozygous missense mutation p.P259L (g.chr9:137242994:C>T (hg38)). Parents were not tested. **(f)** P6 is patient recruited for whole exome sequencing for genetic diagnosis of PCD, followed by targeted analysis of variants in *TUBB4B* (NM_006088.6) which identified a heterozygous *de novo* missense variant [c.776C>T; p.P259L] in exon 4. Both the parents were also tested and confirmed to be wild type (WT) at this locus. **(g)** Summary of variants identified in all patients (P1-P6) as heterozygous for c.776C>T, p.P259L missense variant in exon 4 (below) compared to control sequence (above). Base sequence, amino-acid sequence and codon numbers are shown. Location of base substitution is underlined and amino-acid substitution is depicted in red. **(h)** WES followed by targeted analysis of variants in *TUBB4B* (NM_006088.6) identified a heterozygous in-frame 30 bp duplication variant [c.723_752dup; p.(F242_R251dup)] in exon 4 in patient P7, that was verified by Sanger sequencing (proband, lower chromatogram, compared to a healthy individual). DNA from the parents of P7 were not available for the segregation analysis.**(i)** Proband P8 was recruited as a duo for WGS which on analysis identified a heterozygous missense mutation affecting the same residue but different nucleotide from P1-6. Here, p.P259S (chr9:137242993:C>T (hg38)) was identified in *TUBB4B* only in the patient, and was not present in the parent. DNA from the other parent was not available for segregation analysis. **(j)** Proband P9 in our study, was recruited as part of a family for WES to diagnose profound sensorineuronal disease (SND) including early onset vision and hearing loss, as well as respiratory disease consistent with PCD. A variant c.1072C>T (p.P358S) in exon 4 of the *TUBB4B* gene (NM_006088.5) was identified in only the patient sample in a heterozygous state, confirming *de novo* inheritance. Results confirmed by Sanger sequencing. **(k)** Proband P10, recruited with a history of bilateral sensorineural hearing loss, blindness (SND), chronic kidney disease with nephromegaly and hypertension, chronic productive cough and a history of recurrent sinus and ear infections consistent with PCD. A variant c.1072C>T (p.P358S) in exon 4 of *TUBB4B* (NM_006088.5) was identified in only the patient sample in a heterozygous state, confirming *de novo* inheritance. **(l)** Patient P11 was recruited with a history of congenital heart disease, specifically restrictive cardiomyopathy, as well as hearing and sight loss (SND), plus PCD features such as severe inner ear infections. WES was performed on the parent, sibling and proband initially, which identified the variant c.1072C>T (p.P358S) in exon 4 of *TUBB4B* (NM_006088.5). Inheritance was confirmed by Sanger sequencing in the clinical genetics lab on the proband and the other parent's samples, where the presence of the variant in the heterozygous state was detected only in the patient. **(m)** Summary of variants identified in all patients (P9-P11) as heterozygous for c.1072C>T, p.P358S missense variant in exon 4 (below) compared to control sequence (above). Base sequence, amino-acid sequence and codon numbers are shown. Location of base substitution is underlined and amino-acid substitution is depicted in red. In all pedigrees, males and females are designated by the squares and circles, respectively. Filled symbols represent affected probands. Red arrow above chromatograms highlights the affected residue.

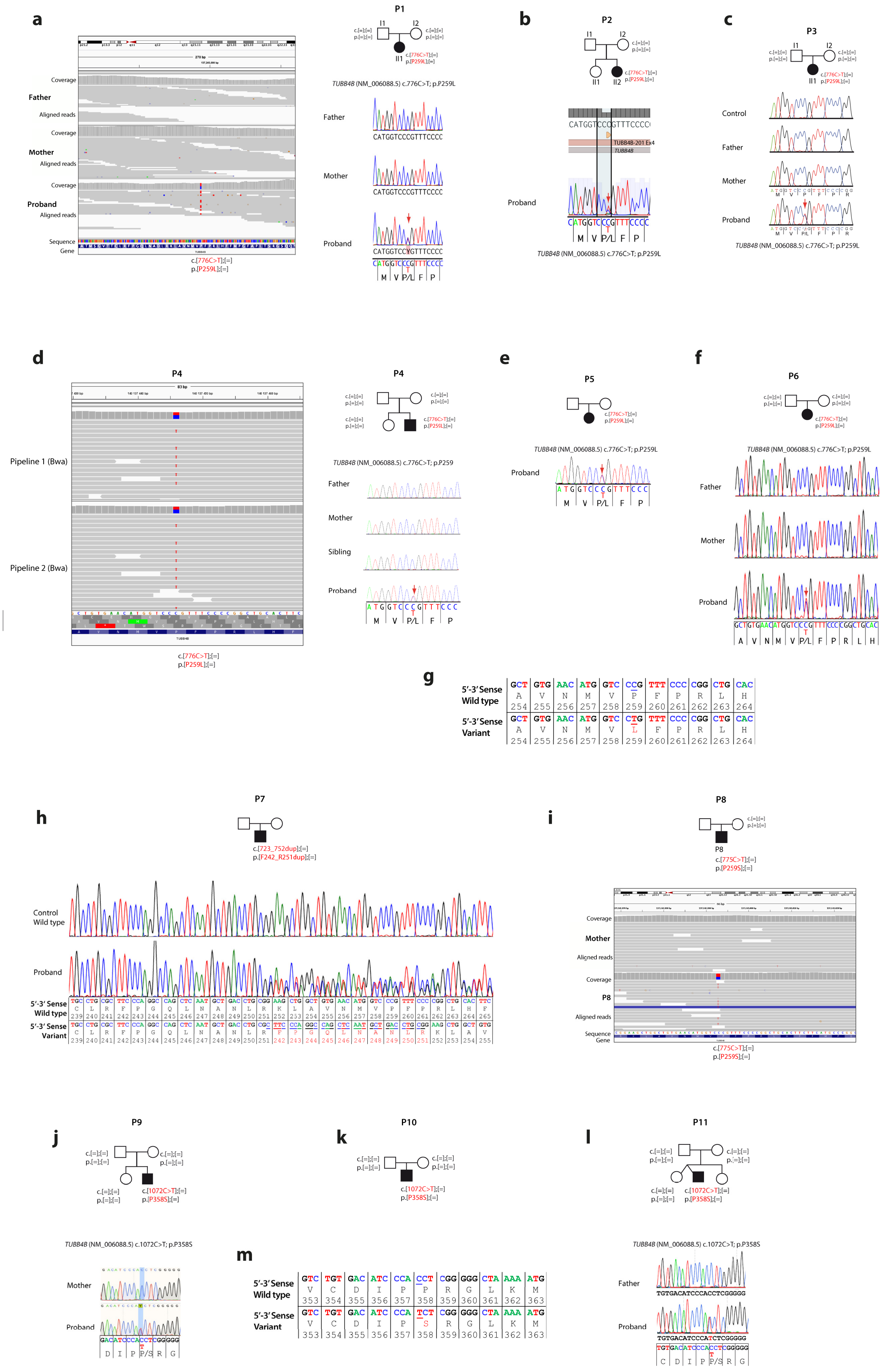
a
P1
Father
Mother
Proband
Coverage
Aligned reads
Sequence
Gene
c.[776C>T];[=]
p.[P259L];[=]
TUBB4B (NM_006088.5) c.776C>T; p.P259L
b
P2
c
P3
Control
d
P4
Pipeline 1 (Bwa)
Pipeline 2 (Bwa)
Sibling
e
P5
f
P6
g
5'-3' Sense Wild type
5'-3' Sense Variant
h
P7
c.[723_752dup];[=]
p.[F242_R251dup];[=]
Control Wild type
i
P8
c.[775C>T];[=]
p.[P259S];[=]
j
P9
c.[1072C>T];[=]
p.[P358S];[=]
TUBB4B (NM_006088.5) c.1072C>T; p.P358S
k
P10
l
P11
m

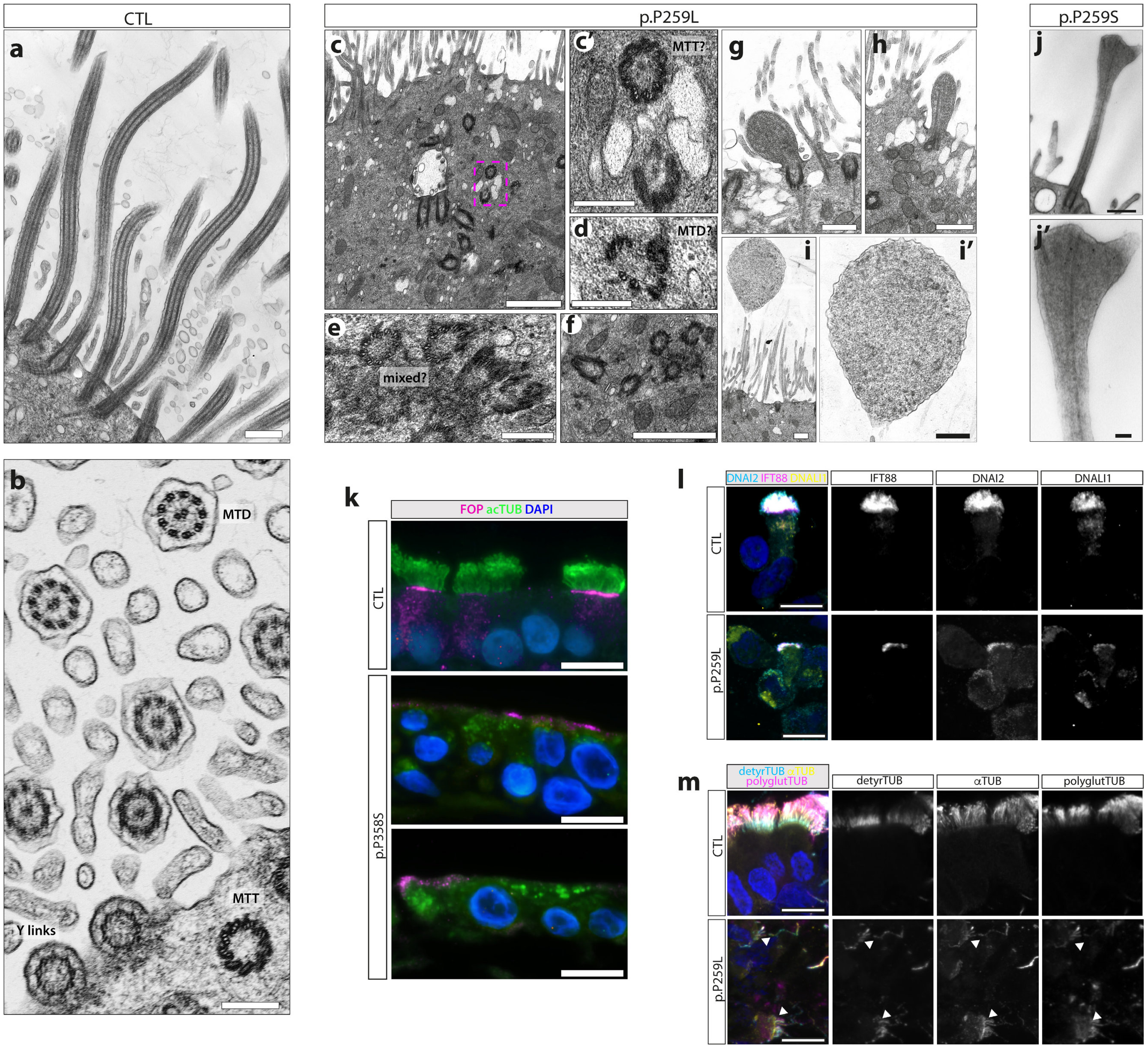


### Extended data- Figure 5: *TUBB4B* variants affect ciliary number, length and microtubule PTMs.

**(a-j')**TEM of nasal brush epithelial cells from control (a,b) and PCD patients with p.P259L variant (P3, c-i') or p.P259S (P8, j,j'). In control cells, dense, long cilia on the surface(a,b), which in cross-section clearly transition through docked basal body (MTDs), transition to MTDs exiting the cell with characteristic Y-links of transition zone and then to '9+2' MTDs of the axoneme. In PCD patient cells evidence of disrupted centriole assembly and amplification is observed including misoriented and internally docked centrioles without axonemes as well as partial structures (c-f). Dashed magenta ROI in (c) with increased zoom to highlight features (c'). (g-i') P3 TEM shows rare short cilia without clear axonemal microtubules (g,h) or dilated cilia tips with disorganized microtubules and granular material (i,i'). (j,j') P8 TEM of short cilia with bulbous head shows splayed microtubules within the expanded tip. **(k)** Nasal brush epithelial cells were cultured and differentiated in ALI from healthy parent (CTL) and syndromic PCD (P9) patient (lower) before sectioning for immunofluorescence confirming reduced centrioles (FOP: magenta) and loss of axonemes, with abnormal acetylated ($\alpha$-tubulin staining (green) mislocalizing within the cytoplasm. **(l,m)** Immunofluorescence of healthy donor (CTL) or patient (P1) nasal brushings stained for (l) cilia outer and inner dynein arm motors (DNAI2: cyan; DNALI1: yellow) of the IFT-B complex (IFT88: magenta;) and (m) microtubule post-translational modifications ($\alpha$-tubulin: yellow; polyglutamylated tubulin: magenta; detyrosinated tubulin: cyan). Scale bars represent: 10 $\mu$m (k-m), 1 $\mu$m (c,f), 500 nm (j, f), 250 nm (c'-e, g,-i'), and 100 nm (j).

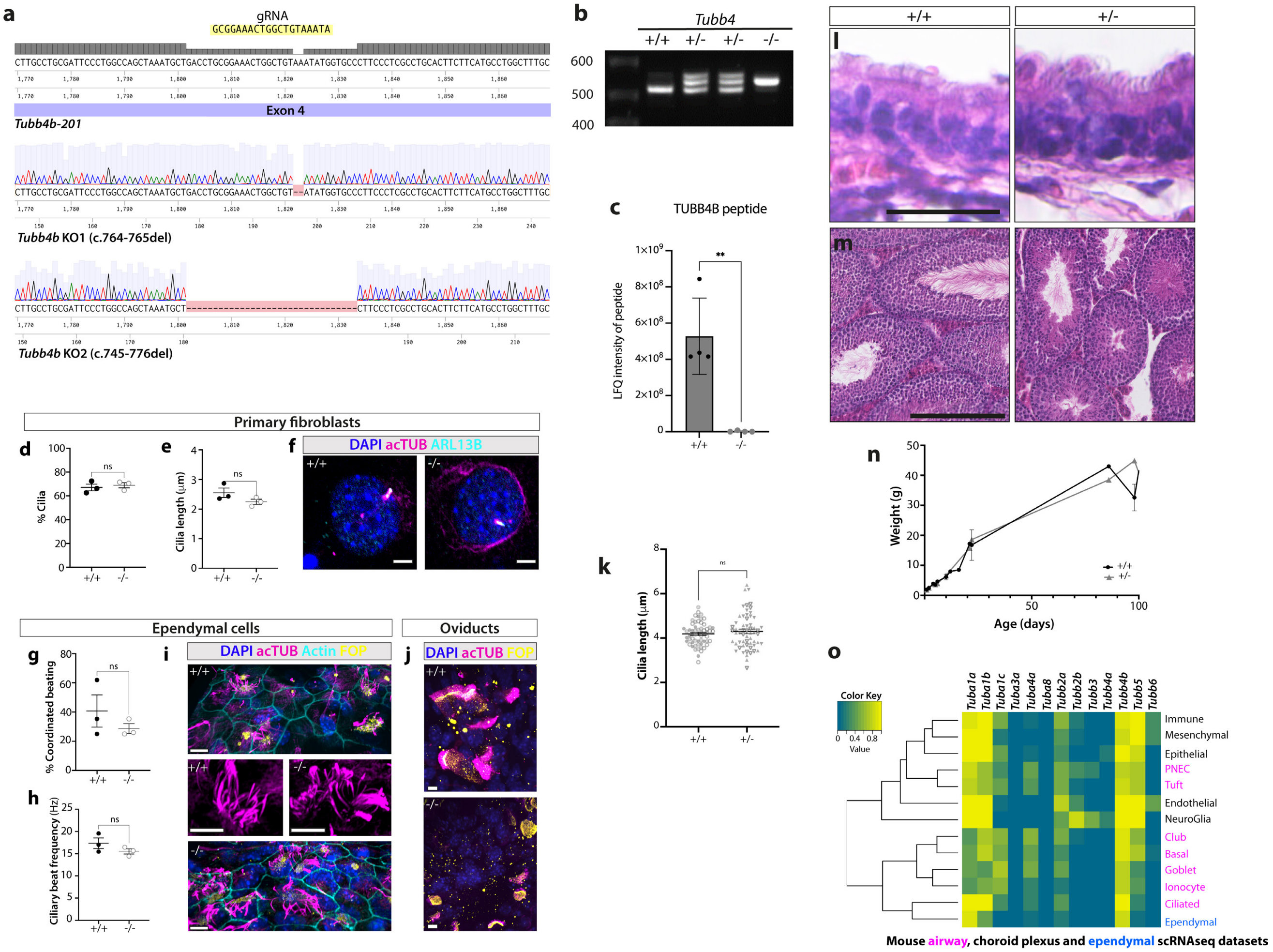


### Extended data- Figure 6: Generation of *Tubb4b* knock-out mice support a dominant negative mechanism of disease for patient pathogenic variants *in vivo* as haploinsufficiency for *Tubb4b* has no effect.

**(a-c)** Two independent deletion alleles were generated by Cas9 genome editing in the 4th exon of *Tubb4b* and founder screening. Both alleles cause a frame shift and premature termination codon. (c) Both alleles are protein null for TUBB4B unique peptides by mass spectrometry (shown for KO2). **(d-f)** *Tubb4b*$^{-/-}$ primary cilia on fibroblasts show no difference in percentage ciliation (d), cilia length (e) or gross structure by immunofluorescence (f). **(g-i)** Despite the pronounced hydrocephaly phenotypes visible *in vivo*, mutant primary ependymal cells differentiated in vitro show no difference by high-speed video microscopy in percentage of cells with coordinated beating (g), ciliary beat frequency (h) or quantification of cilia structures stained for immunofluorescence (i). **(j)** Whole-mount immunofluorescence of oviducts reveals a similar arrest of ciliogenesis as in airway epithelia. **(k-n)** Heterozygous KO animals (shown for KO2) are phenotypically normal, showing normal trachea cilia numbers (k) and lengths (l), normal fertility, as shown by normal spermatogenesis (m), and postnatal survival/growth (n). **(0)** Single cell RNASeq heatmaps show the proportions of cells expressing each $\alpha$- and $\beta$-tubulin isotype (RNA count greater than zero) in each cell type identified in the population from published mouse lung (magenta)(68), ependymal (blue)(66) and choroid plexus (black)(67) single cell RNASeq datasets. Rows are clustered on similarity of proportions among cell types (scale 0-1). Cilia length and number of ciliated cells in fibroblasts were quantified using ARL13B as a marker, N = 3 biological replicates per genotype with (h) n >139 cells per biological replicate and (i) n > 87 cells per biological replicate, mean values are plotted. Ependymal beat coordination and beat (k, l) were calculated from N = 3 biological replicates per genotype, n > 17 cilia measurements per replicate. (j, m, n) Fibroblast Growth Factor Receptor 1 Oncogene Partner (FOP): yellow, acetylated $\alpha$-tubulin: magenta, actin: cyan (m) or ARL13B: cyan (j). Scale bars represent: 250 $\mu$m (m), 25 $\mu$m (l), 10 $\mu$m (i, j) and 5 $\mu$m (f). (c, k, n) Graphic bars represent the mean ± SEM derived from N=4 biological replicates (c), N=3 biological replicates, n>18 cells/sample (k) and N= 155 animals. (d, e, g, h) Statistical analyses were carried out by the PLSD Fisher test. Graphic bars represent the mean ± SEM derived from three biological replicates. Student's t-test: ns, not significant; **, p $\leq$0.01.

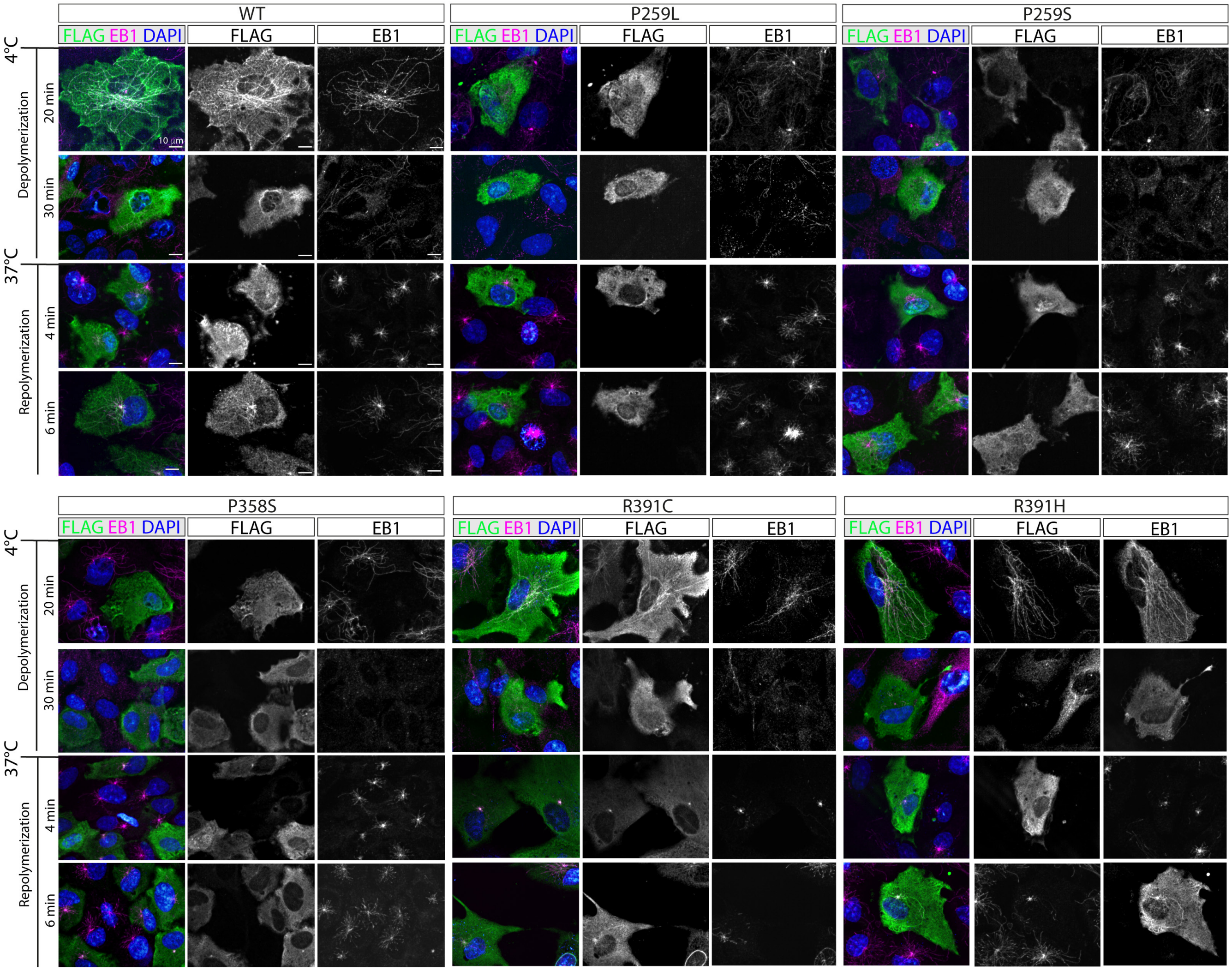


### Extended data- Figure 7: Disease-causing TUBB4B variants alter microtubule dynamics.

Microtubule network dynamics analysis of RPE1 cells overexpressing TUBB4B variants, showing immunostaining of FLAG-tagged TUBB4B (green) and EB1 (magenta) protein upon cold-induced depolymerization (20 and 30 minutes) and repolymerization at 37 °C (4 and 6 minutes). See **Figure 3(g, h)** for quantification of repolymerization. Scale bars represent: 10 $\mu$m.

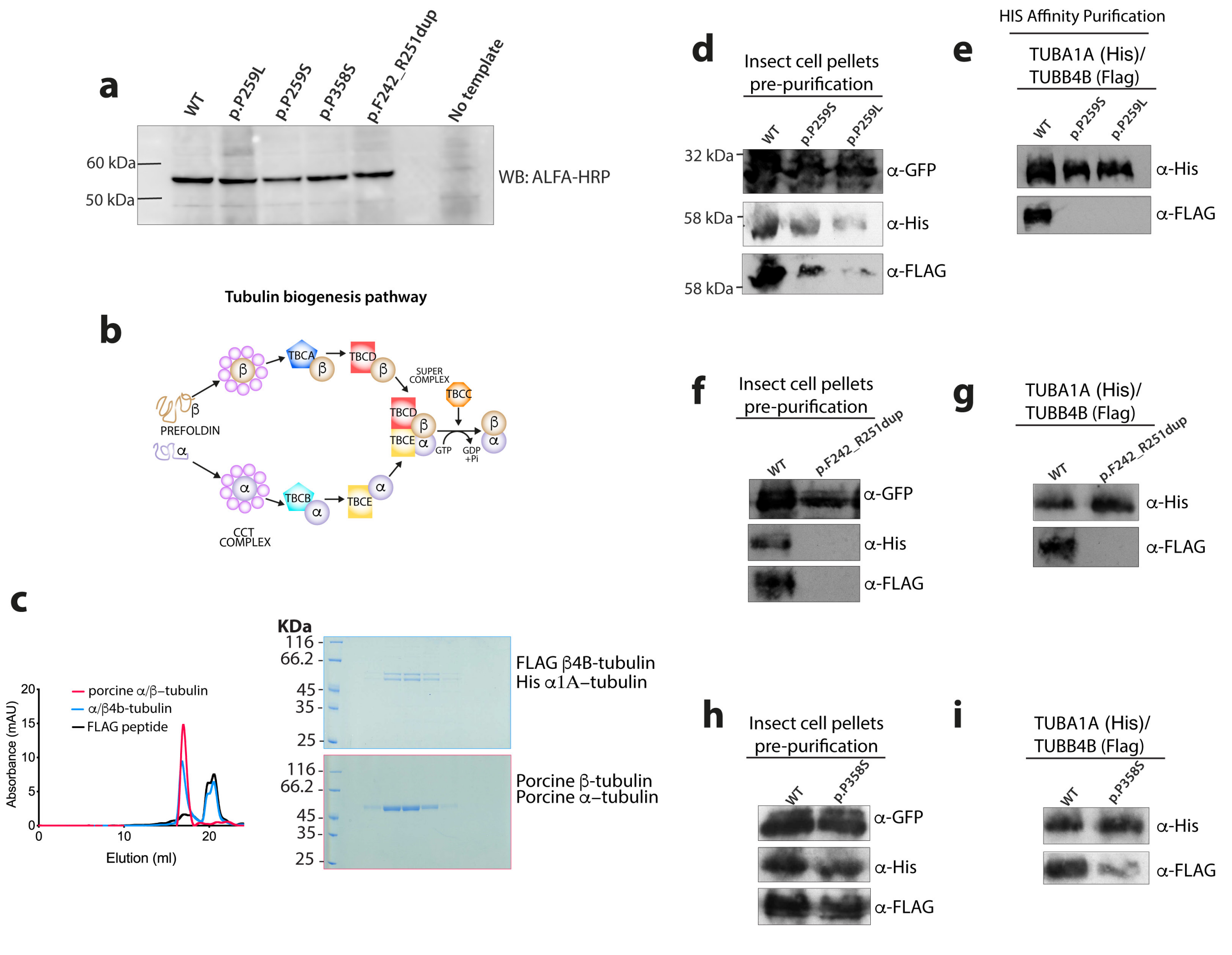


### Extended data- Figure 8: *TUBB4B* variants cause dominant negative disease through distinct molecular mechanisms affecting heterodimerization or polymerization.

**(a)** Immunoblot of *in vitro* translated ALFA-tagged control and *TUBB4B* patient variants. All PCD-only (p.P259L, p.P259S, Dup) and syndromic PCD+SND p.P358S TUBB4B variants are stable. **(b)** Schematic of tubulin heterodimer assembly pathway. Binding of partially folded tubulin molecules as they emerge from the ribosomes by prefoldin and subsequent folding by the cytoplasmic chaperonin CCT. These quasi-native tubulin intermediates interact with five tubulin-specific chaperones named tubulin cofactors A through E (TBCA–TBCE). The native assembly-competent tubulin is released from a supercomplex that contains both $\alpha$- and $\beta$-tubulin and cofactors C–E, upon hydrolysis of GTP by $\beta$-tubulin in the supercomplex. **(c-i)** Effects of *TUBB4B* variants on tubulin stability and heterodimerization were investigated using Sf9 insect cells expressing recombinant bacmids containing human tubulin-$\alpha$1A with an internal His tag and human tubulin-$\beta$4B-FLAG. The recombinant bacmid also contains a GFP reporter gene to monitor infection of Sf9 cells, and is used as a loading control. (c) Size exclusion chromatography analysis and elution profile for the indicated constructs for porcine $\alpha/\beta$-tubulin (red), FLAG-eluted tubulin-$\alpha$/tubulin-$\beta$4B (blue) and FLAG peptide (black) (c, left panel). Coomassie-stained gels showing elution profiles for the corresponding protein complexes (c, right panel). Whole insect cell pellets were analyzed by immunoblot probed with anti-His and anti-FLAG antibodies for $\alpha$-tubulin and $\beta$-tubulin respectively to monitor expression (p.P259L/S (d); Dup (f); p.P358S (h)), as well as GFP to monitor transduction. After elution from His-tag affinity Ni-NTA beads, immunoblots for recombinant tubulin heterodimers were probed with anti-His and anti-FLAG antibodies for $\alpha$-tubulin and $\beta$-tubulin respectively to monitor heterodimerization (p.P259L/S (e); Dup (g); p.P358S (i)).

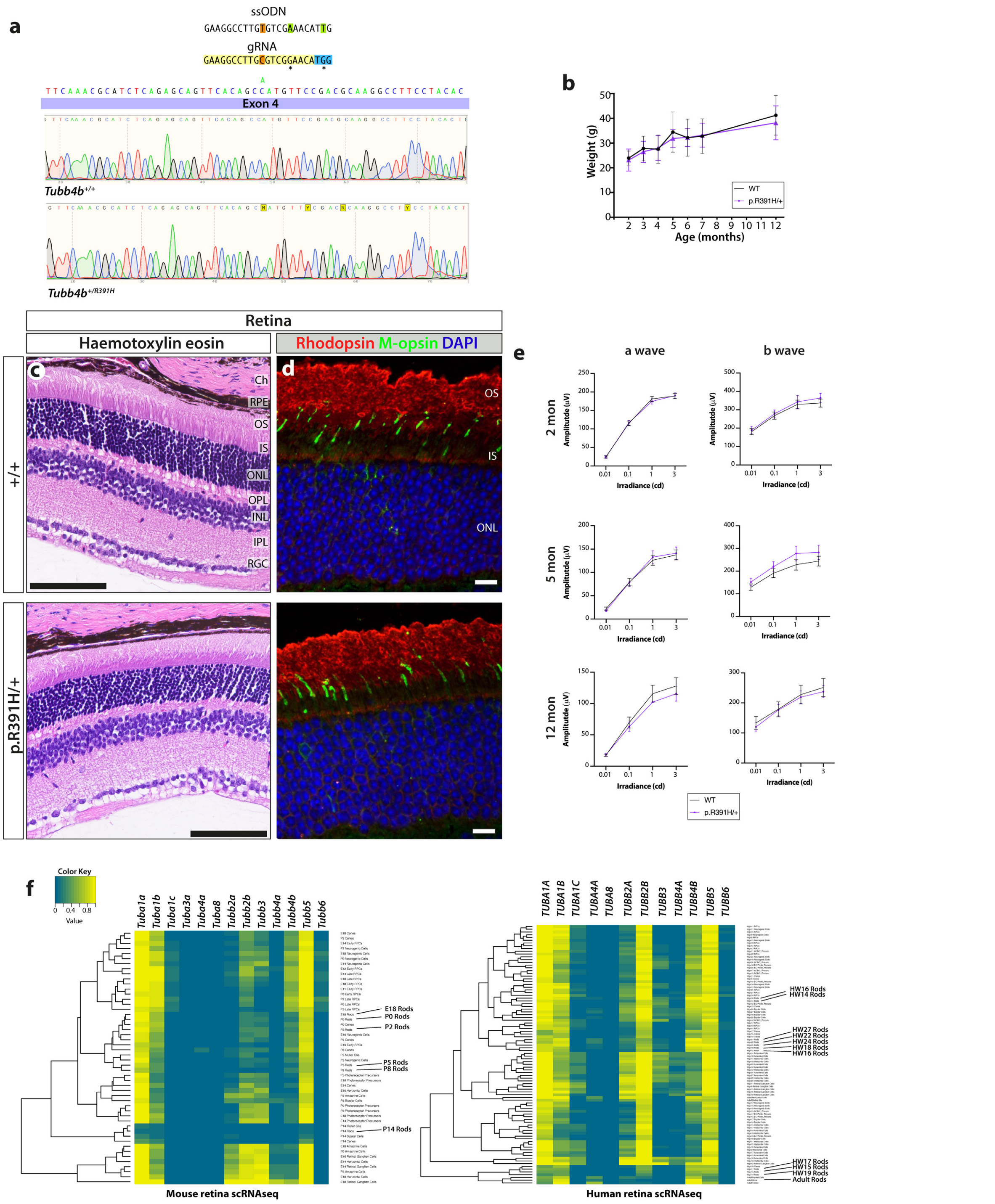


### Extended data- Figure 9: Engineered point mutations in *Tubb4b* do not recapitulate patient phenotypes in the mouse retina.

**(a)** Strategy to generate LCA KI p.R391H allele by Cas9 genome editing in the 4th exon of *Tubb4b* and Sanger sequencing to validate. **(b)** $Tubb4b^{R391H/+}$ animals show no decrease in fitness or survival postnatally. **(c-e)** $Tubb4b^{R391H/+}$ animals show no early onset or age-related degeneration of photoreceptors by histology (c) or immunofluorescence (d) (rhodopsin: red; M-opsin: green) in 4 month adult mice. (e) $Tubb4b^{R391H/+}$ animals show no evidence of physiological changes in neuroretina with age as shown by electroretinogram unlike the p.R391H/+ human patients. **(f)** Species-specific differences in expression of TUBB4B in the neuroretina are likely to account for these differences. Heatmaps show the proportions of cells expressing each tubulin (RNA count greater than zero) in each cell type identified in the population from published mouse (left)(35) and human (right)(34) neuroretina single cell RNASeq datasets. Rows are clustered on similarity of proportions among cell types (scale 0-1). Scale bars represent: 100 $\mu$m (c) and 10 $\mu$m (d). (b, e) Graphic bars represent the mean ± SEM derived from N>3 animals per time point.

**Table 4.1 CRISPR guides and repair templates**

| Name | Sequence | Application | Source |
|---|---|---|---|
| *Tubb4b* Exon4 p.R391H guide | 5' - GAAGGCCTTGTGTCGAAACATTG - 3' | CRISPR guide for generation of *Tubb4b* $^{R391H/+}$ mouse | GeneArt Precision gRNA synthesis kit (ThermoFisher Scientific, USA) |
| *Tubb4b* Exon4 p.R391H patient variant KI ssODN | 5' - GAAGGCCTTGCGTCGGAACATGG - 3' | Repair template for generation of *Tubb4b* $^{R391H/+}$ mouse | |
| *Tubb4b* Exon4 p.P259L guide | 5' - CCATATTTACAGCCAGTTTCCGC - 3' | CRISPR guide for generation of *Tubb4b* $^{P259L/+}$ mouse | GeneArt Precision gRNA synthesis kit (ThermoFisher Scientific, USA) |
| *Tubb4b* Exon4 p.P259L patient variant KI ssODN | 5'- GCTGGGTGAGCTCAGGAACTGTCAGGGCACGGTACTGCTGGCTGCCCCGGCTGGTCAAGGGGGCAAAGCCAGGCATGAAG AAGTGCAGGCGAGGGAAGaGCACCATgTTaACgGCCAGTTTCCGCAGGTCAGCATTTAGCTGGCCAGGGAATCGCAGGCAAG TGGTTACCCCACTCATGGTGGCGGACACTAGATGGTTC - 3' | Repair template for generation of *Tubb4b* $^{P259L/+}$ mouse | IDT |
| *Tubb4b* Exon4 p.P259L patient silent mutation ssODN | 5' - GCTGGGTGAGCTCAGGAACTGTCAGGGCACGGTACTGCTGGCTGCCCCGGCTGGTCAAGGGGGCAAAGCCAGGCATGAAG AAGTGCAGGCGAGGGAAGGgCACCATgTTaACgGCCAGTTTCCGCAGGTCAGCATTTAGCTGGCCAGGGAATCGCAGGCAAG TGGTTACCCCACTCATGGTGGCGGACACTAGATGGTTC - 3' | Repair template for generation of *Tubb4b* $^{+/+}$ mouse: silent mutation control | IDT |
| *Tubb4b* Exon4 p.P259L guide | 5' - AAATGCTGACCTGCGGAAAC - 3' | CRISPR guide for generation of *Tubb4b* $^{P259L/+}$ mouse | GeneArt Precision gRNA synthesis kit (ThermoFisher Scientific, USA) |
| *Tubb4b* Exon4 p.P259L patient variant KI ssODN | 5'- CACCCACTTACGGTGACCTGAACCATCTAGTGTCCGCCACCATGAGTGGGGTAACCACTTGCCTGCGATTCCCTGGCCAGCTA AATGCTGATCTCCGCAAGCTGGCTGTAAATATGGTGCTCTTCCCTCGCCTGCACTTCTTCATGCCTGGCTTTGCCCCCTTGACCA GCCGGGGCAGCCAGCAGTACCGTGCCCTGACA - 3' | Repair template for generation of *Tubb4b* $^{P259L/+}$ mouse | IDT |
| *Tubb4b* Exon4 p.P259S guide | 5' - CCATATTTACAGCCAGTTTCCGC - 3' | CRISPR guide for generation of *Tubb4b* $^{P259S/+}$ mouse | GeneArt Precision gRNA synthesis kit (ThermoFisher Scientific, USA) |
| *Tubb4b* Exon4 p.P259S patient variant KI ssODN | 5'- GCTGGGTGAGCTCAGGAACTGTCAGGGCACGGTACTGCTGGCTGCCCCGGCTGGTCAAGGGGGCAAAGCCAGGCATGAAG AAGTGCAGGCGAGGGAAGGaCACCATgTTaACgGCCAGTTTCCGCAGGTCAGCATTTAGCTGGCCAGGGAATCGCAGGCAAG TGGTTACCCCACTCATGGTGGCGGACACTAGATGGTTC - 3' | Repair template for generation of *Tubb4b* $^{P259S/+}$ mouse | IDT |
| *Tubb4b* Exon4 p.P259S patient silent mutation ssODN | 5' - GCTGGGTGAGCTCAGGAACTGTCAGGGCACGGTACTGCTGGCTGCCCCGGCTGGTCAAGGGGGCAAAGCCAGGCATGAAG AAGTGCAGGCGAGGGAAGGgCACCATgTTaACgGCCAGTTTCCGCAGGTCAGCATTTAGCTGGCCAGGGAATCGCAGGCAAG TGGTTACCCCACTCATGGTGGCGGACACTAGATGGTTC - 3' | Repair template for generation of *Tubb4b* $^{+/+}$ mouse: silent mutation control | IDT |
| *Tubb4b* Exon4 p.P358S guide | 5' - GCCGACATTTTCAGGCCCCG - 3' | CRISPR guide for generation of *Tubb4b* $^{P358S/+}$ mouse | GeneArt Precision gRNA synthesis kit (ThermoFisher Scientific, USA) |
| *Tubb4b* Exon4 p.P358S patient variant KI ssODN | 5'- GTCGGAACATGGCTGTGAACTGCTCTGAGATGCGTTTGAACAGCTCCTGAATAGCGGTGCTGTTGCCAATGAAGGTGGCCGA CATTTTCAGGCCCCGAGATGGAATGTCACAGACAGCTGTCTTCACATTGTTGGGGATCCACTCAACAAAGTAGCTGCTGTTCTT GTTTTGGACATTAAGCATCTGTTCGTCCACCTCC - 3' | Repair template for generation of *Tubb4b* $^{P358S/+}$ mouse | IDT |
| *Tubb4b* Exon4 p.P358S patient silent mutation ssODN | 5' - GTCGGAACATGGCTGTGAACTGCTCTGAGATGCGTTTGAACAGCTCCTGAATAGCGGTGCTGTTGCCAATGAAGGTGGCCGA CATTTTgAGcCCgCGAGgTGGAATGTCACAGACAGCTGTCTTCACATTGTTGGGGATCCACTCAACAAAGTAGCTGCTGTTCTTG TTTTGGACATTAAGCATCTGTTCGTCCACCTCC - 3' | Repair template for generation of *Tubb4b* $^{+/+}$ mouse: silent mutation control | IDT |
| *Tubb4b* Exon4 KO guide | 5' - CCATATTTACAGCCAGTTTCCGC - 3' | CRISPR guide for generation of *Tubb4b* $^{+/-}$ mouse, generated while targeting p.P259L and p.P259S mutations | GeneArt Precision gRNA synthesis kit (ThermoFisher Scientific, USA) |

**Table 4. 2 Site mutagenesis and genotyping primers.**

**Site mutagenesis primers**

| Patient variant | Primer name | Primer Sequence | Process |
|---|---|---|---|
| **P259L** | p.P259L_forward | 5' CTGTGAACATGGTCCTGTTTCCCCGGCTGCA 3' | Site directed mutagenesis |
| | p.P259L_reverse | 5' TGCAGCCGGGGAAACAGGACCATGTTCACAG 3' | |
| **P259S** | p.P259S_forward | 5' GCTGTGAACATGGTCTCGTTTCCCCGGCTGC 3' | Site directed mutagenesis |
| | p.P259S_reverse | 5' GCAGCCGGGGAAACGAGACCATGTTCACAGC 3' | |
| **P358S** | p.P358S_forward | 5' CTGTCTGTGACATCCCATCTCGGGGGCTAAAAAT 3' | Site directed mutagenesis |
| | p.P358S_reverse | 5' ATTTTTAGCCCCCGAGATGGGATGTCACAGACAG 3' | |
| **R391C** | p.R391C_forward | 5' CACGGCCATGTTCCGGTGCAAGGCCTTCCTGCAC 3' | Site directed mutagenesis |
| | p.R391C_reverse | 5' GTGCAGGAAGGCCTTGCACCGGAACATGGCCGTG 3' | |
| **R391H** | p.R391H_forward | 5' CACGGCCATGTTCCGGCACAAGGCCTTCCTGCAC 3' | Site directed mutagenesis |
| | p.R391H_reverse | 5' GTGCAGGAAGGCCTTGTGCCGGAACATGGCCGTG 3' | |

**Genotyping primers**

| Mouse Line | Primer name | Primer Sequence | Process |
|---|---|---|---|
| *Tubb4b* $^{R391H}$ | Tubb4b_R391H F | 5' CTGAAAATGTCGGCCACCT 3' | PCR followed by Sanger sequencing |
| | Tubb4b_R391H R | 5' GACTAAGACAGCTCTAAGCC 3' | |
| *Tubb4b* $^{KO}$ | Tubb4b KO F | 5' GTTGAGCCCTACAATGCCAC 3' | PCR followed by Sanger sequencing |
| | Tubb4b KO R | 5' GAAGGTGGCCGACATTTTCA 3' | |

**Extended data- Figure 10: List of guides, repair ssODNs, primers and antibodies used in this study.**

**Table 4.3 Primary antibodies**

| Antigen | Antibody/Clone Name | Host species | Source | Application |
|---|---|---|---|---|
| α-tubulin | DM1A | Mouse | Sigma | WB (1:1000); IF (1:2000, MeOH) |
| α-tubulin | YL1/2, ab6160 | Rat | Abcam | WB (1:2000); IF (1:1000, PFA) |
| Acetylated α-tubulin | 6-11B-1, T6793 | Mouse | Sigma | IF (1:1000-1:5000) |
| ALFA-HRP | N1505-HRP | Camelid nanobody | Nanotag | WB (1:1000) |
| ARL13B | 17711-1-AP | Rabbit | ARL13B | IF (1:1000, PFA) |
| Centrin | 20H5 04-1624 | Mouse | Merck | IF (1:500, MeOH w. PE or PFA) |
| Detyrosinated tubulin | AA12, ab254154 | Mouse | Abcam | IF (1:1000, PFA) |
| DNALI1 | HPA028305 | Rabbit | Sigma | IF (1:200, PFA) |
| DNALI1 | N-13 | Goat | Santa Cruz | IF (1:75, PFA) |
| DNAH5 | HPA035364 | Rabbit | Sigma | IF (1:200, PFA) |
| DNAI2 | IC8 | Mouse | Abnova | IF(1:100, PFA) |
| EB1 | 5/EB1 | Mouse | BD Biosciences | IF (1:200, MeOH) |
| FLAG | ab95045 | Goat | Abcam | IF (1:5000, MeOH) |
| FLAG | M2, F1804 | Mouse | Sigma | WB (1:1000) |
| FOP | 11343-1-AP | Rabbit | Proteintech Group | IF (1:500, PFA) |
| Gamma tubulin | GTU88, T6557 | Mouse | Sigma | IF (1:500, MeOH w PE) |
| GFP | MMS-118P | Mouse | Covance | WB (1:5000) |
| His | 02-10667 | Rabbit | RayBiotech | WB (1:1000) |
| IFT88 | 13967-1-AP | Rabbit | Proteintech Group | IF (1:100, PFA) |
| M-Opsin | OSR00222W | Rabbit | ThermoFIsher Scientific | IF (1:500, PFA) |
| Polyglutamylated tubulin | GT335, AG-20B-0020-C100 | Mouse | AdipoGen Life Sciences | IF (1:1000, PFA or MeOH) |
| Pericentrin | ab4448 | Rabbit | Abcam | IF (1:1000, MeOH) |
| Rhodopsin | 4D2, NBP2-59690 | Mouse | Novus Biologicals | IF (1:500, PFA) |
| TBCD | 14867-1-AP | Rabbit | Proteintech Group | WB (1:1000) |

| Probes | Cat number | Modification | Source | Application |
|---|---|---|---|---|
| ActinRed™ 555 ReadyProbes™ | R37112 | Rhodamine | Thermo Fisher Scientific | IF (2 drops/mL) |
| Alexa Fluor 647 Phalloidin | A22287 | Alexa-647 | Thermo Fisher Scientific | IF (1:500) |

**Table 4.4 Secondary antibodies**

| Antigen | Host Species | Dilution | Source | Application |
|---|---|---|---|---|
| ECL α-Mouse IgG, HRP-conjugated | Sheep | 1:7500 | GE Healthcare UK Ltd | WB |
| ECL α-Rabbit IgG, HRP-conjugated | Sheep | 1:7500 | GE Healthcare UK Ltd | WB |
| HRP-conjugated α-Rabbit IgG H + L | Goat | 1:5000 | BioRad | WB |
| HRP-conjugated α-Mouse IgG H + L | Goat | 1:5000 | BioRad | WB |
| Alexa 488, 568, 594, 647-conjugated-α-Mouse | Donkey | 1:500 | Invitrogen Molecular Probes | IF |
| Alexa 488, 594, 647 -conjugated-α-Rabbit | Donkey | 1:500 | Invitrogen Molecular Probes | IF |
| Alexa 555, 647-conjugated-α-Goat | Donkey | 1:500 | Invitrogen Molecular Probes | IF |
